# Systematic review and mega-analysis of the peripheral blood transcriptome in depression implicates dysregulation of lymphoid cells and histones

**DOI:** 10.1101/2025.05.01.25326802

**Authors:** Chaitanya Erady, Richard Bethlehem, Ed Bullmore, Mary-Ellen Lynall

## Abstract

**Background:** Depression has been associated with transcriptomic changes in peripheral blood. However, the contribution of specific immune cell subsets or pathways remains unclear, and findings have been variable across previous studies, which have not tended to account for sample cellular composition.

**Methods:** We performed a systematic review of peripheral blood transcriptome studies in depression. For the five datasets meeting criteria (total N=6,011), we performed harmonized reprocessing and cell-composition-adjusted differential gene and transcript analyses, followed by a bias- and inflation-adjusted weighted Z-score mega-analysis. We investigated the biological pathways and cell subsets implicated by the results. We also performed a sex-stratified gene network mega-analysis using consensus weighted gene co-expression network analysis (WGCNA).

**Results:** Few genes showed robust differential gene expression (DGE) in depression. Depression was reproducibly associated with decreases in replication-dependent histones, and with a decrease in oxidative phosphorylation pathways in females only. Cell source analyses implicated lymphoid cells (T cells and NK cells) as likely contributors to the depression differential expression signature. WGCNA mega-analysis revealed multiple consensus modules associated with depression, with a *PUF60*-related module upregulated in both female and male depression in sex-stratified analyses. Two genes predicted to be causally relevant to depression by transcriptome-wide association studies (*GPX4* and *GYPE*) showed significant DGE.

**Conclusions:** These results are convergent with immunogenetic evidence implicating lymphoid cell dysregulation in depression, while also highlighting histone alterations as a key molecular signature in depression. They also indicate the importance of large-scale datasets for biomarker discovery in the context of heterogeneous disorders like depression.

## Introduction

Depression is a debilitating disorder of the mind and body that results from a complex interplay of genetic, environmental, psychological and social factors (1). There is increasing evidence for the involvement of the immune system in the pathogenesis of major depressive disorder (MDD), at least in some patients (2). Depression has been associated with increases in acute phase reactants and pro-inflammatory cytokines as well as abnormalities in peripheral immune cells (3), and randomized trial data suggest a beneficial effect of some immunotherapeutics on depressive symptoms (4). Analyses of the genetic risk variants associated with depression suggest a causal role for immunity in depression. For example, levels of the cytokine IL-6 have been estimated to increase risk of depression using Mendelian randomization (5), and depression risk variants are enriched at epigenetically active sites in lymphoid immune cells, especially activated T cells (6). If we are to capitalize on these findings to move towards personalized treatments, it is critical that we better understand the cellular immunopathology of depression, and find reproducible biomarkers of immune dysfunction that can be detected in patients.

To this end, multiple studies have examined blood-derived gene expression in depression. A previous meta-analysis in 2,521 participants (the largest to date) found 342 genes to be differentially expressed in depression (7). However, this analysis did not account for cellular composition, so genes detected may reflect the known effects of depression on immune cell counts in peripheral blood (3, 8), as acknowledged by the authors. Moreover, previous work has not harmonized processing of datasets using individual-level raw data (‘mega-analysis’) or corrected for the p-value inflation that affects genome-wide molecular epidemiology studies (9), nor acknowledged the importance of sex in immunopathogenesis (10). Previous sex-stratified analyses of MDD-associated differential expression from bulk brain samples have found significant differences between males and females (11). However, analyses at the single nucleus level have highlighted that the pattern of cell-intrinsic MDD-associated changes in brain is broadly consistent between males and females (12). Whether there are robust sex-specific immune-related transcriptomic changes in MDD remains unclear.

A complementary approach to univariate (gene-based) differential expression analysis is to test for disease-associated differences in groups (modules) of genes that tend to be coregulated e.g. using weighted gene co-expression network analysis (WGCNA). Previous work has highlighted innate immune co-expression modules as important in MDD (7, 13); but subsequent work has shown that without correction for cellular composition, most co-regulatory links detected simply reflect variation in cell composition rather than cell-intrinsic regulatory networks (14).

Here, we present the largest blood-based depression case-control transcriptomic mega-analysis to date (2,375 cases and 3,606 controls), applying harmonized gene- and transcript-level analyses while accounting for differences in sample cellular composition (see **Figure S1** for overview). We used both sex-pooled and sex-stratified approaches to test for MDD-associated changes in expression and compared these to genetically predicted expression profiles. To identify the cellular processes, immune cell subsets, and co-expression networks implicated in depression, we applied pathway enrichment, cell origin analysis, and consensus weighted gene co-expression network analysis (WGCNA) across all datasets, correcting for cell composition throughout. We also specifically tested for dysregulation in pathways previously implicated in MDD, including T cell activation, mitochondrial function, ribosomes, and histones. Summary statistics, along with (where permitted) harmonized processed individual-level data, are provided as a resource.

## Methods and Materials

### MDD case-control study selection

A systematic literature review was conducted following the Preferred Reporting Items for Systematic Reviews and Meta-Analyses (PRISMA) guidelines. The primary inclusion criteria were blood gene expression datasets (RNA-Seq or microarray) from MDD case-control studies in an adult population. Studies published up until August 2021 were considered for inclusion and study selection was performed by two reviewers independently (C.E. and M.E.L.) using the covidence.org platform (see **Supplementary Methods**). Five publications met criteria; one of the five analyzed two relevant MDD case-control datasets; access for one of these was possible via direct contact with the authors. Five MDD case-control datasets - three using RNA-Seq (BIODEP, Le, and Mostafavi) and two using microarray (dbGaP and HiTDiP) - met criteria and were included in this analysis (**Table 1**). No ethical approval was required; this is a secondary analysis of published studies complying with ethical standards.

**Table 1.** Sample information for the five MDD case-control datasets evaluated for transcriptomic dysregulation. Datasets included in the mega-analysis comprise both whole blood and peripheral blood mononuclear cell (PBMC) samples, assayed using either RNA-sequencing (RNA-seq) or microarray. For further information on included datasets see **Supplementary Text** and **Table S2**. Ancestries are reported as per original publications; NA indicates information was not available.^1^For Le, ancestry was noted as Caucasian 76%; African-American 7.6%; Native American 2.5%; native Hawaiian/Pacific Islander 1.3%; Asian American 2.5%; Other 9.5%. ^2^For dbGaP, current depression information was available for 97% of the MDD samples. CNT, Control; MDD, Major depressive disorder; MAD, median absolute deviation; bps, base pair; poly(A), polyadenylated.

| Dataset | Tissue type | RNA expression assay method | Total n | n (CTL) | n (MDD) | % Female | Median age (± MAD) | Ancestry | Currently depressed |
| --- | --- | --- | --- | --- | --- | --- | --- | --- | --- |
| BIODEP | PBMC | RNA-seq (75bps paired-end reads, poly(A) selected) | 211 | 47 | 164 | 69.2 | 33.8 ± 9.2 | UK | 75.6% |
| Le | PBMC | RNA-seq (150bps paired-end reads, poly(A) selected) | 153 | 78 | 75 | 58.2 | 28 ± 8.9 | Mixed<sup>1</sup> | NA |
| Mostafavi | Whole blood | RNA-seq (50bps single-end reads, poly(A) selected) | 896 | 444 | 452 | 70.3 | 46 ± 13.3 | Caucasian | 44.7% |
| dbGaP | Whole blood | Microarray (Affymetrix U219 array plate) | 4540 | 2976 | 1564 | 66.1 | 35 ± 11.9 | European | 37.1%<sup>2</sup> |
| HiTDiP | Whole blood | Microarray (Affymetrix U133 plus 2.0 array plate) | 181 | 61 | 120 | 74.0 | 52.9 ± 13.5 | UK | NA |
| Total N = |  |  | 5,981 | 3,606 | 2,375 |  |  |  |  |

### Differential expression analyses

We first performed per-dataset harmonized processing, followed by per-dataset differential gene expression (DGE), differential transcript expression (DTE) and differential transcript usage (DTU) analysis. The gene expression sequencing platforms, sample isolation procedures, and available covariates differed between the five studies (**Table 1**). In brief (see **Figure S2** and **Supplementary Text**), RNA-seq reads were aligned to GRCh38 v84 with STAR (v2.5.2b) (15); gene counts were generated from uniquely mapped reads using featureCounts (v.1.5.1) (16) and transcripts quantified with salmon (v1.4.0) (17). Counts were transformed with limma-voom, incorporating voomWithQualityWeights (18). For microarrays, dbGaP-processed counts were used; otherwise, .CEL files were normalized with affy (v1.68.0) robust multi-array average (RMA) method (19). Gene expression was modelled using limma (v.3.46.0) (20).

We aimed to correct for the variable cell subset composition across samples. Our in-house dataset (BIODEP) assayed cell counts by flow cytometry; for other datasets, cell count enrichment scores were estimated from gene expression data using xCell (v1.1.0) (see **Supplementary Text**). For each included dataset, the DGE and DTE ‘main model’ applied was:

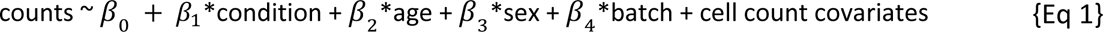

‘Cell count covariates’ refers to assayed or estimated counts of B cells, CD4+ T cells, CD8+ T cells, NK cells and monocytes for PBMC samples; the ‘main model’ [Eq 1] for whole blood samples additionally included neutrophil and eosinophil counts as cell count covariates, but not basophils (due to the poor correlation of xCell-estimated basophils with recorded basophils, **Figure S4**). In addition to this main model, per-dataset sensitivity DGE analyses included (a) the main model without cell count covariates; (b) the main model with BMI as an additional covariate, for the three datasets where BMI was available (BIODEP, Mostafavi, and dbGaP); and, (c) for datasets using whole blood samples (Mostafavi, dbGaP, and HiTDiP), the main model with an additional term for (xCell-)estimated platelet counts (see **Supplementary Methods**).

Three of the five MDD case-control datasets were RNA-Seq data, permitting additional transcript-level mega-analysis. Per-dataset DTU analysis was performed using DEXSeq (v1.36.0) (21) followed by stageR (v1.12.0) (22) for gene-wise correction of transcript p*-*values. See **Figure S2** and **Supplementary Text** for details of DGE/DTU pipelines.

### Mega-analysis of depression-associated expression

Per-dataset gene and transcript differential expression results were meta-analyzed using a bias- and inflation-corrected weighted Z-score method. Sample size-weighted Z score combination methods are recommended over effect size combination methods when different gene expression platforms are included in a meta-analysis (23, 24). Bias and inflation represent the deviation of the test statistics’ mean and variance, respectively, from the theoretical null distribution. For each dataset (*i*), Z-scores (calculated from DGE P-values with direction of effect taken from the sign of DGE log_2_FC) were corrected for inflation and bias using R package bacon (v1.18.0) (9). These modified Z-scores (*x_i_*) were combined over the *n* datasets to produce a meta-analytic weighted Z-score as follows:

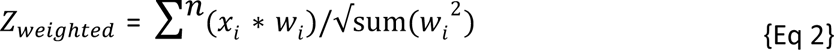

Where *W_i_* is the effective sample size for dataset *i* calculated as:

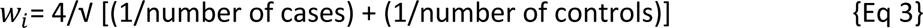

P-values corresponding to the meta-analytic Z-scores were corrected for multiple testing using the BH method with FDR<0.05 considered significant. Mega-analysis was not applied to DTU results as the assumptions for weighted Z-score meta-analysis are not met (non-uniform distribution of p-values under the null hypothesis).

### Enrichment analysis

To determine the pathways dysregulated in MDD, gene set enrichment analyses (GSEA) (25) was applied to the mega-analytic DGE/DTE results. GSEA was performed on genes ranked by signed DGE/DTE p-values, using clusterProfiler GSEA function (v3.18.1, n=10,000 permutations) (26) with BH FDR correction. For DTE, where multiple transcripts mapped to the same gene, the gene with the most significant p-value was retained. DGE GSEA used pathway gene lists from the expert-curated Reactome database v75 (27) and the experimentally-derived ImmuneSigDB database (28), which contains 4,872 gene sets identified from 389 immunological studies. DTE GSEA used Reactome pathways only, as transcript expression is less interpretable with respect to ImmuneSigDB, which is based on gene-level experimental results.

We also used GSEA to directly test for dysregulation of T cell activation in MDD by re-analyzing a human *ex vivo* dataset (29) that profiled transcriptomic responses of naive and memory CD4+ T cells stimulated with varying levels of anti-CD28 and anti-TCR. Using the same methods as ImmuneSigDB (see **Supplementary Text**), we identified the genes most strongly up- and down-regulated by activation and compiled them into pathway lists. We tested whether these activation-responsive pathways were enriched in our MDD-associated DGE results.

### Cell source analysis

The MDD-associated DGE signature may arise from gene expression changes in one or more of the cell types present in blood. To identify which cell subsets are likely contributing to the bulk DGE signal, we used LRCell, a computational tool that estimates cell-type contributions by regressing a DGE signature against marker profiles of known cell types (30). We applied LRCell to the MDD DGE signature using a multimodal PBMC single-cell dataset (31) as the reference.

### Concordance of genetically-predicted and observed MDD-associated gene expression

We aimed to test whether the genes predicted to be causally important for MDD can be detected as dysregulated in peripheral blood samples. To compare our observed DGE results to genetically-predicted DGE (TWAS), we drew on a recent multi-ancestry GWAS meta-analysis (32) which identified 169 loci associated with MDD and conducted a TWAS on these results using FUSION. We obtained the blood-only TWAS results (based on SNP weights from GTEx v8) from the authors. In blood, 60 genes were genetically predicted to be associated with MDD at the authors’ TWAS-wide significance level (P < 1.37 x 10^-6^). To capture the similarity and divergence of the TWAS and observed gene expression profiles (including non-genome wide significant genes), we conducted a threshold-free rank-rank hypergeometric overlap (RRHO) analysis using the R package RRHO2 (v1.0) (33).

### Mega-analysis of dysregulation of core cellular processes

We also tested for concerted up- or down-regulation of groups of functionally related genes involved in core cellular processes previously linked to MDD. We tested whether the median log_2_FC of each gene group differed between MDD and controls using a two-sided permutation test, performed separately for pooled, female, and male samples. To control for expression-related biases, replacement genes were sampled without replacement from a background gene set matched for baseline expression (within ±0.1 log_2_CPM). Median log_2_FCs from n=1,000 matched permutations were compared to the observed median log_2_FC to calculate p-values, which were then meta-analyzed across datasets using the weighted Z-score method (as above) with BH multiple comparisons adjustment. Because of the computational intensity of this method, we focused on 13 curated groups of genes previously implicated in MDD and expected to change expression as a group: ribosomal genes, mitochondrial genes, nuclear-encoded mitochondrial genes, components of the OXPHOS electron transport chain, replication-dependent and -independent histones, and cell cycle genes associated with G1, G1/S, S, G2, G2/M, and M phases (see **Supplement** for gene set curation and further details of permutation testing).

### Consensus weighted gene co-expression network analysis (WGCNA)

In addition to testing for dysregulation of groups of genes based on biological similarity, we also sought to identify data-driven modules of co-expressed genes consistently dysregulated in MDD, while accounting for sample cellular composition. We used consensus WGCNA (34) to identify separately male and female gene co-expression networks consistently present across multiple datasets (see also **Supplementary Methods**). Log expression values for all 5 datasets were corrected for study center, age, and cell counts and filtered to retain only genes common to all datasets (*n* = 11,857). Consensus gene-gene similarities were identified by first calculating signed topological overlap matrices (TOMs) for each dataset based on gene-gene correlations calculated using biweight midcorrelation (bicor). TOMs were combined across datasets such that each element of the consensus TOM matrix was set as the minimum of that element across all datasets; this conservative method retains only gene-gene connections present in all datasets. Hierarchical clustering of the dissimilarity matrix (1-TOM) was used to construct gene network dendrograms and detect modules using the WGCNA R package (v1.70-3)(35) with soft-thresholding powers of 12 for female and 10 for male samples based on scale-free topology and mean connectivity (further details in **Supplement**).

To characterize the consensus modules detected, we calculated module eigengenes, hub genes and enrichment of biological processes. Per-module biological process gene ontology (GO) term over-representation analyses were performed using a one-sided Fisher exact test via clusterProfiler enrichGO to select module-associated terms with BH FDR<0.05; the significant GO term with the highest gene ratio was selected as the module representative term. Eigengene networks were constructed and module preservation scores calculated (36) to assess inter-dataset similarity in consensus networks (see **Supplement**).

## Results

### Study selection and characteristics

Two researchers independently reviewed the literature for MDD case-control transcriptomic datasets (see **Figure S3** PRISMA diagram and **Supplement**); of 424 PubMed results, 56 studies were selected for full-text review. The 5 studies that met inclusion criteria were used in our analyses (**Table 1**). These comprise a mixture of bulk RNA-Seq (n=3) and microarray (n=2) datasets isolated from PBMCs (n=2) and whole blood (n=3). We refer to these as BIODEP (37), Le (38), Mostafavi (39), dbGaP (40) and HiTDiP (13). Where known (**Table 1** and **Table S1**), most datasets included both currently depressed and remitted MDD patients, were majority female, majority European ancestry with high concurrent antidepressant use (see also **Supplementary Text**). Few studies (BIODEP and a subset of dbGaP samples) had sufficiently granular cell count data (e.g. T, B cells) to support deconvolution of gene expression data. Thus, bioinformatic deconvolution (via xCell) was used to infer cell count enrichments from gene expression data. Estimated cell enrichments showed moderate correlations with assayed cell counts where these were available (**Figure S4**), except basophils, which were thus excluded from subsequent transcriptomic models. Assayed and estimated cell counts showed significant case-control differences (**Table S2**), further motivating correction for cellular composition in transcriptomic models.

### Gene-level mega-analysis

Individual dataset results for DGE were obtained (Eq. 1) and meta-analyzed using a bacon-adjusted weighted Z-score analysis (Eq. 2) for genes present in all five datasets (see **Methods**). Inclusion of cell counts reduced Z-score inflation from 1.31 to 1.21 (**Figure S5**). For DTE, the same meta-analysis model was applied to the transcripts detectable in all three RNAseq datasets. There was little evidence of robust DGE: two genes were under-expressed in MDD in the sex-pooled DGE meta-analysis (*ACRBP* and *CDIPT*), with 38 genes additionally DGE in the female-only meta-analysis, and no significant DGE in the male-only meta-analysis (see **Figure 1A**). In a traditional meta-analysis (without correction for bias and p-value inflation), 124 genes were called as upregulated and 73 genes as downregulated in MDD.

**Figure 1.**
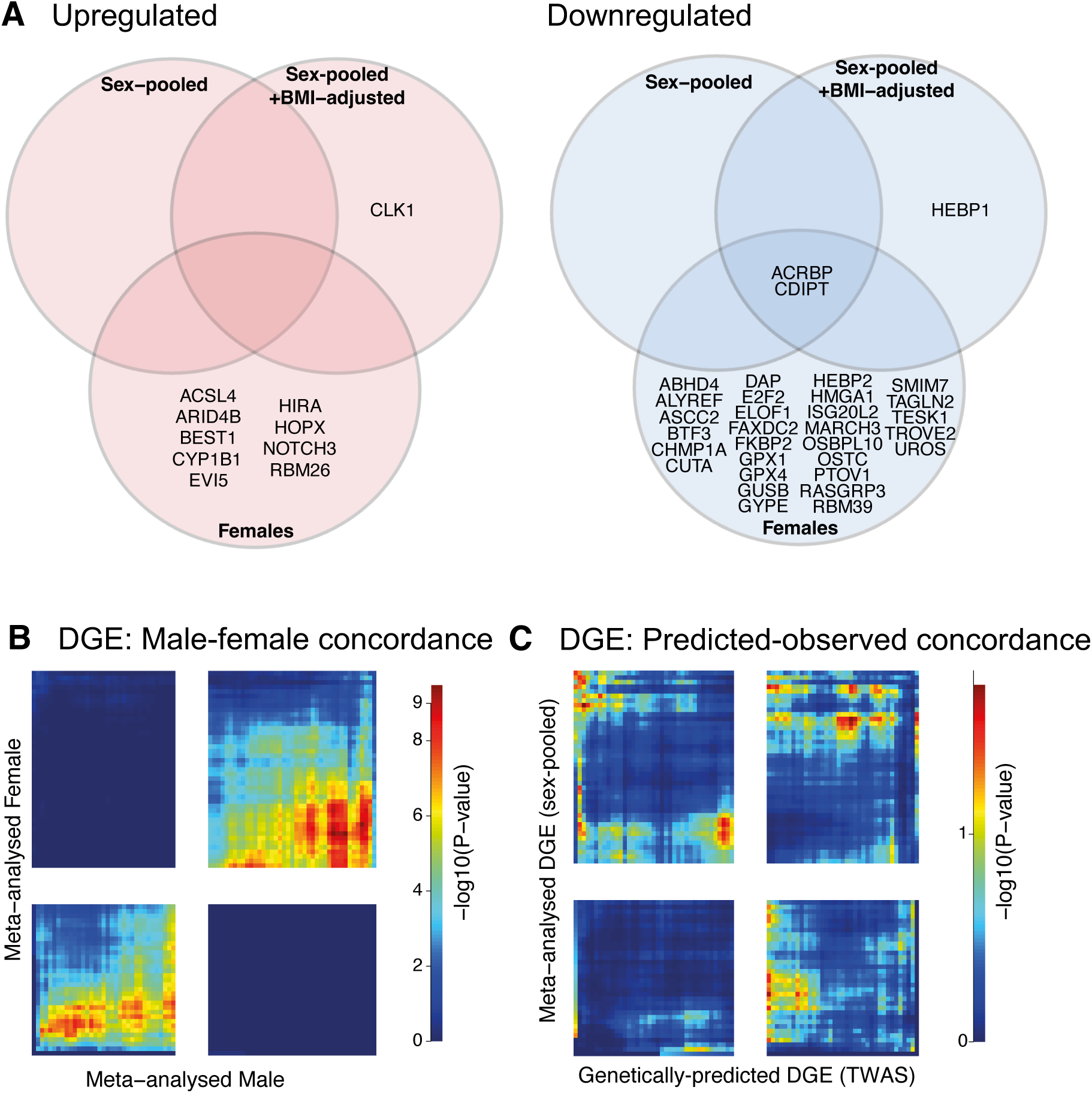
Concordance between observed vs genetically-predicted differential gene expression (DGE) (A) Venn diagrams illustrate overlap among significantly differentially expressed genes (FDR<0.05) identified in sex-pooled, sex-pooled BMI-adjusted, and female-specific meta-analyses. No genes were differentially expressed at FDR<0.05 in the male-specific meta-analysis. (B) Rank-rank hypergeometric overlap (RRHO) heatmap showing significant concordance between male and female meta-analyzed DGE results (overall Pearson’s r = 0.095, P < 2.2e-16). On the axes, genes from each list (male DGE and female DGE) are ordered from most significantly upregulated (left of x axis for males, bottom of y axis for females) to most significantly downregulated (right, top). Each heatmap pixel represents the significance (-log10 hypergeometric p-values) of overlap between genes ranked above a certain threshold in both lists. Strong concordance of gene expression changes is shown by signal in the top-right (downregulated) and bottom-left (upregulated) quadrants (red indicating stronger concordance), and discordance shown in the opposite quadrants. (C) RRHO heatmap comparing observed meta-analyzed DGE results with genetically predicted DGE (TWAS, Meng et al., 2024). There was poor concordance between real-world expression differences and genetic predictions (note differing heatmap scale), and overall, no significant correlation between observed and predicted DGE (Pearson’s r = 0.0015, P = 0.91).

Examining DGE signals without thresholding at a genome-wide significance level, results from the sex-stratified meta-analyses were significantly correlated between males and females (r = 0.095, P < 2.2e-16) with significant male-female concordance in the pattern of differential expression (**Figure 1B**); we also observed cross-dataset concordance in DGE signal that did not cluster either by tissue type or by assay technology (**Figure S6**). Given the low overall DGE signal, and the male-female concordance, we focused further investigation on the sex-pooled DGE analysis.

We examined whether DGE genes (FDR < 0.2) were also associated with differential protein abundance in a recent UK Biobank MDD study (41). ARID4B, LILRB1, and IGSF8 showed concordant upregulation at the protein level in MDD (FDR < 0.05).

### Concordance between genetically predicted and observed differential gene expression

To test to what extent our observed pattern of DGE results matched the predicted effects of MDD genetic risk variants, we tested the concordance between our observed DGE results and genetically predicted MDD-associated DGE from a recent depression TWAS using blood eQTL data (32). Of 1,550 TWAS-significant genes (P < 0.05), only two - *GPX4* and *GYPE* - showed concordant observed downregulation in females with MDD in our meta-analysis (FDR < 0.05), with similar trends (raw P<0.05) in the pooled analysis (see **Supplement**). Overall, there was no correlation between observed and genetically predicted DGE (r = 0.0015, P = 0.91), and little concordance in the threshold-free pattern of DGE (**Figure 1C**), even after re-analysis without cell composition correction to more closely match the TWAS methodology (**Figure S6C**).

### Pathway analysis of DGE signature

To characterize the biology of the meta-analytic DGE signature, we used GSEA to test for enrichment of curated human pathways (Reactome) and gene sets from immunological studies (ImmuneSigDB). In MDD, there was significant downregulation of the mitochondrial complex 1 biogenesis pathway (NES= -1.883, FDR = 0.019) (a result driven by females; **Figure 2A**). MDD was also associated with downregulation of pathways related to cell cycle control including G1/S DNA damage checkpoints (NES= -1.877, FDR = 0.012) and SCF-mediated degradation of the cyclin dependent kinase inhibitors (NES= -1.893, FDR = 0.012); results which showed the same direction of effect in males and females (**Figure 2A**). For the sex-stratified meta-analyses, pathway enrichment analyses additionally highlighted some sex-divergent pathways, including a female MDD-specific downregulation of mitochondrial electron transport chain related pathways and female MDD-specific upregulation of PPAR-ɑ signaling (**Figure S7**). Given previous genetic evidence implicating dysregulated CD4+ T cell activation in depression (6), we also tested for evidence of altered T cell activation, generating up- and down-regulated gene sets from a human experimental study of naive and memory CD4+ T cell activation under different stimulation conditions (29). There was a downregulation of T cell activation signatures in the meta-analytic MDD signature, for both genes normally downregulated, and genes normally upregulated upon T cell activation (**Figure 2B**).

**Figure 2.**
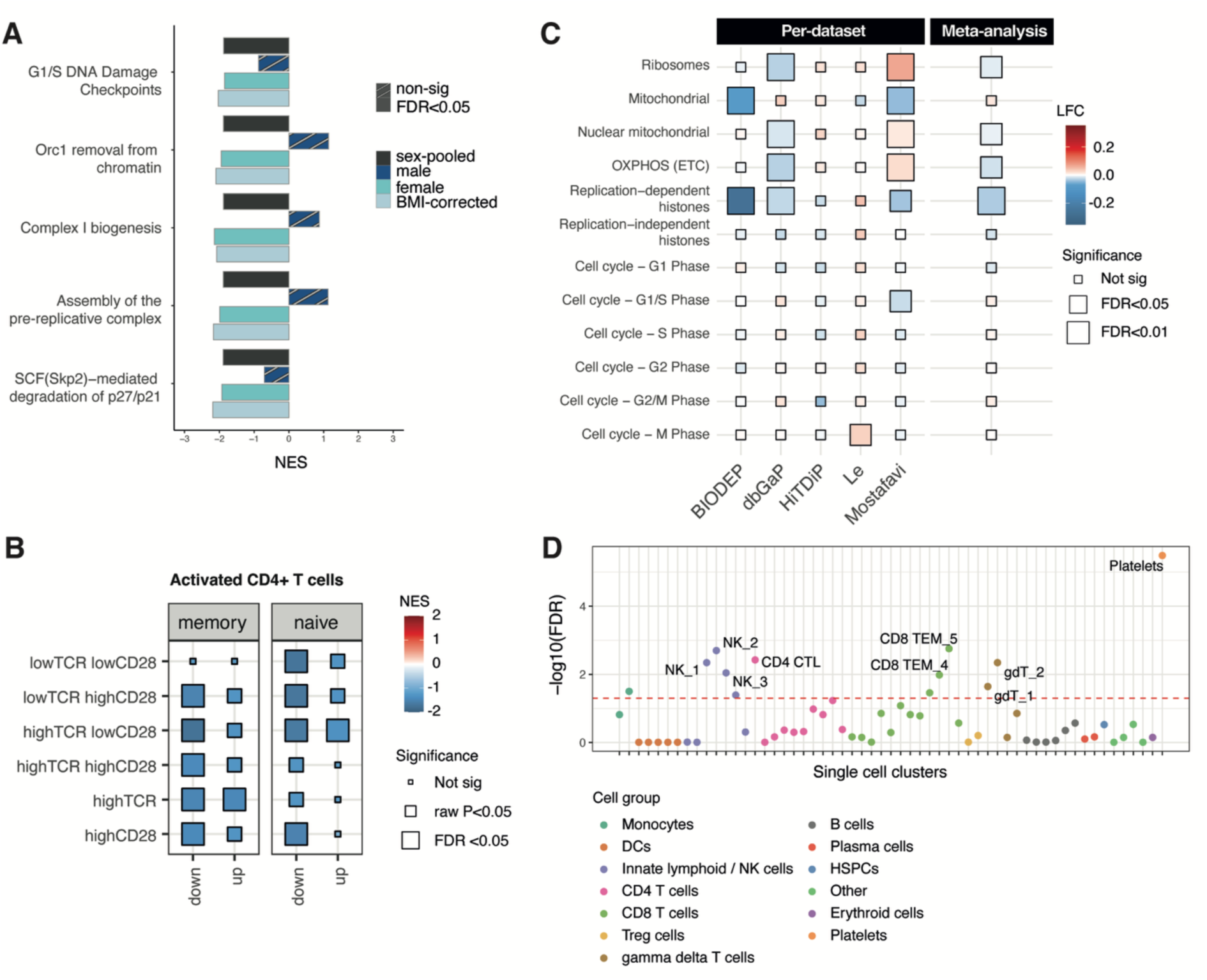
Pathways and cell subsets implicated by differential gene expression analysis. (A) Pathway or gene set enrichment analysis (GSEA) was performed on signed p-values obtained from differential gene expression (DGE) mega-analysis of pooled samples. For pathways with FDR<0.05 in the sex-pooled mega-analysis, the top 5 most downregulated Reactome pathways (by normalized enrichment score, NES, y-axis) are shown; no Reactome pathways were significantly upregulated. NES indicates enrichment scores normalized for differences in gene set sizes. Enrichments for the sex-pooled meta-analysis are shown by dark grey bars; male-only meta-analysis by dark blue bars; female-only meta-analysis by blue bars; and BMI-corrected sex-pooled meta-analysis by pale blue bars. Stripped bars indicate non-significant enrichments (FDR > 0.05). No ImmuneSigDB pathways were significantly differentially expressed. (B) Enrichment of T cell activation pathways in depression DGE signature by GSEA. Data represent gene sets derived from CD4+ T cell stimulation experiments using varied concentrations of anti-CD28 and anti-T cell receptor (TCR) to activate naive and memory T cells. Tile color indicates normalized enrichment score for the T cell activation gene set in the MDD depression signature; ‘down’ and ‘up’ indicate the gene sets down- and upregulated upon activation (compared to rest) in CD4+ T cells; tile size indicates FDR p-values (C) MDD-associated changes in median expression of genes involved in core cellular processes for pooled samples are shown. A permutation test was conducted (where genes with similar expression were used as a replacements) to calculate p-values, which were corrected for multiple testing using the BH method. Size of rectangles indicates these FDR values. For each dataset, tile color indicates median MDD-associated LFC of genes present in a specific gene group. The meta-analytic tile color shows the mean of these LFCs across datasets, weighted by sample size. Per-dataset permutation values were meta-analyzed using the weighted Z-score method. ETC, electron transport chain; G, Gap or growth phase; S, Synthesis phase; M, Mitotic phase. See **Figure S8** for sex-stratified results. (D) Cell origin analysis indicates the cell subsets estimated to be driving the MDD-associated expression signature. Dot plot shows regression of the MDD expression signature (sex-pooled analysis) against cell subset marker genes derived from a peripheral blood single cell dataset (see **Methods**). X-axis shows the immune cell types tested, with legend showing the major cell subgroups. Y-axis shows the significance of each cell type enrichment by -log10(FDR); dashed red line indicates FDR=0.05; the most significant enrichments are labelled. See also **Figure S9**.

### Dysregulation of core biological processes

We assessed whether gene sets corresponding to core cellular processes previously implicated in depression showed coordinated expression changes using permutation testing (**Methods; Figure 2C**). Results showed downregulation of replication-dependent histones (meta-analytic FDR=0.001), but not replication-independent histones (FDR=0.31) in MDD. Sex-stratified meta-analyses showed significant effects of MDD on replication-dependent histones in females (FDR=0.009) and a similar trend in males (FDR=0.1) (**Figure S8**). The MDD-associated decrease in replication-dependent histones was observed in both microarray datasets (polyA-selected) and RNA-seq datasets, suggesting that an MDD-associated decrease in canonical polyA(-) replication-dependent histones is not the primary driver of this signal. Supporting this, there was no overall change in the expression of genes enriched at different stages of the cell cycle in MDD (**Figure 2C**). Additionally, significant decreases were found in nuclear mitochondrial genes and electron transport chain (OXPHOS) genes in MDD (both FDR=0.04), driven mainly by females (**Figure S8**).

### Cellular origin analysis

To identify the likely cellular source of depression-related gene expression changes, we used LRCell to regress the threshold-free DGE profile (already corrected for cellular composition) against single-cell marker profiles from PBMCs (31). This analysis implicated intrinsic changes in CD8+ T effector memory (TEM) cells (coef=0.006, FDR=0.002), NK cell subsets (particularly NK2 i.e. CD16+CD38+ NK cells; coef=0.007, FDR=0.002; **Figure 2D**) and platelets (coef=0.05, FDR<0.001). Several key genes contributed to signals in CD8+ TEM and NK cells, including *CD247*, *GZMH*, *IL2RG*, and *CD7*.

The platelet-associated signal might reflect differences in platelet counts between MDD cases and controls rather than intrinsic platelet gene expression changes. Assayed platelet counts were unavailable for any whole blood samples so were not included in the main meta-analytic model. We thus conducted sensitivity analyses using xCell-estimated platelet counts, which significantly differed between MDD and controls in two datasets (dbGaP, p<0.001; HiTDiP, p=0.017). Adjusting for these estimates in the meta-analysis eliminated the platelet signal but preserved the CD8+ TEM and NK cell signals (**Figure S9**).

### Sensitivity analyses

We performed a BMI-adjusted sensitivity meta-analysis, in which most of the genes and cell subsets prioritized remained significant despite the smaller sample size with BMI available - see **Supplement**. Since the meta-analysis was weighted by sample size, results are likely driven by the largest dataset, dbGaP. Leave one out (LOO) meta-analyses confirmed that dbGaP was the major driver of the individual gene results (**Supplementary Text**).

### Differential transcript expression and differential transcript usage

For the datasets for which transcript-level information was available (RNA-seq datasets), we also performed a DTE meta-analysis (see **Figures S1, S2**). No genes were DTE at a meta-analytic level, likely due to the lower sample size (N=1,260 for DTE vs N=5,981 for DGE) and higher number of multiple comparisons corrected for (38,877 transcripts for DTE vs. 11,845 genes for DGE). Similar to the main meta-analysis, DTE pathway enrichment analyses highlighted decreases in pathways related to cell cycling (G1/S-specific transcription, NES=-2.01, FDR=0.02), but additionally detected increases in multiple pathways suggesting increased translation initiation, in both males and females (see **Supplementary Text**). In the complementary DTU analysis, based on changes in transcript proportions for each gene, 0 genes were DTU in more than one dataset. The non-uniform p-value distribution of DTU results precludes weighted Z-score meta-analysis (see **Supplement**).

### Network meta-analysis using consensus weighted gene co-expression network analysis (WGCNA)

To complement gene-level analyses, we performed consensus WGCNA to identify co-expressed gene modules across all five datasets, analyzing males and females separately due to known sex differences in co-expression (29, 30). We identified 8 consensus modules in females and 11 in males (**Table S3, Figures 3A, 3B**). Module eigengenes summarized expression patterns, and eigengene networks showed high preservation across datasets and platforms (**Supplementary Text, Table S4**). Most genes, however, did not form consensus modules, and few differentially expressed genes from gene-level meta-analyses were module members (**Figure S10**). GO enrichment (**Table S5**) and hub gene identification via kME scores provided biological interpretation.

**Figure 3.**
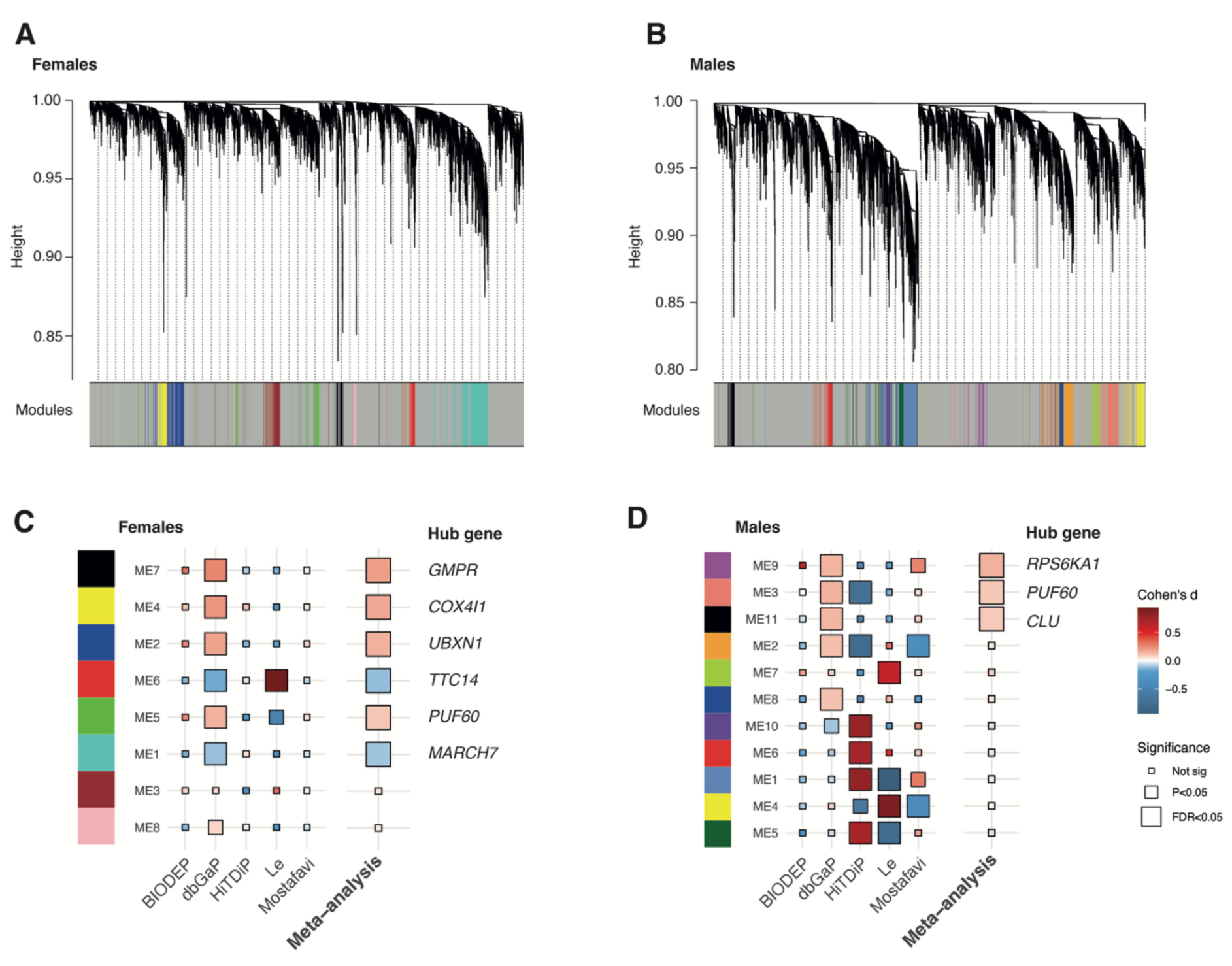
Meta-analysis of consensus gene co-expression modules in MDD. WGCNA dendrograms and modules representing co-expressed genes were identified in pooled CTL and MDD samples for (A) females and (B) males. Dendrogram height (a measure of how similar modules are) is indicated on y-axes (dissimilar modules connect at a larger height - closer to 1.00 on the plot). The modules corresponding to these dendrograms are shown on x-axes, with arbitrary colors used consistently for each module and grey representing genes not assigned to consensus modules. (C, D) Tile plots show WGCNA consensus modules differentially expressed (DE) in MDD vs. controls calculated using a t.test in (C) females and (D) males. Plots are ordered from most significantly DE (top) to least significantly DE (bottom). Plots show module names on the x axis, dataset on the y-axis. Tile color indicates Cohen’s d effect size of DE. Tile size indicates t.test significance. Meta-analysis column tiles show Cohen’s d values weighted by sample size, and the significance level for the weighted z-score meta-analysis of per-dataset results. Tile colors to the left of the module correspond to the module colors below the dendrograms in (A) and (B). Hub gene indicates the genes in each modules with the highest kME i.e. the most connected genes in the module. Sample sizes used in this analysis are as follows: CNT + MDD females = 3,967 samples; CNT + MDD males = 1,966 samples; BIODEP = 205; Le = 150; Mostafavi = 885; dbGaP = 4,515; HiTDiP = 178 samples.

We tested if consensus modules differed between MDD and controls using t-tests in each dataset followed by weighted z-score meta-analysis of these results. In females, 6 of 8 modules, and in males, 3 of 11 modules showed differential expression (**Figures 3C, 3D, Table S5**). In both sexes, modules with hub gene PUF60 were consistently upregulated in MDD. However, these module results were primarily driven by the largest dataset, dbGaP (**Table S6**).

## Discussion

### Altered lymphocyte, mitochondrial and histone pathways in depression

In this first mega-analysis of peripheral blood gene expression in depression, we identified subtle but robust changes in core cellular and immune processes in MDD. Our results particularly implicate altered T cell and natural killer (NK) cell function, supporting previous predictions from immunogenetic analyses (6). Analyses also implicate mitochondrial gene dysregulation (in females only), and a concerted decrease in histones in MDD in both sexes.

Our analyses consistently highlighted dysregulation of lymphocyte function, particularly involving T cells and NK cells. Pathway analysis revealed decreased expression of T cell activation genes in depression. Cellular origin analysis, integrating our results with single-cell data, confirmed NK cells and T cells as the likely sources of the MDD transcriptional signature. One meta-analytically downregulated gene, *CDIPT*, is essential for CD8+ T cell effector function (42). These results align with previous immunogenetic findings (6), which causally implicate lymphoid cells and T cell activation in MDD. These results are interesting in the light of cell count meta-analyses which have demonstrated increased circulating CD4+ T cells, activated T cells, and NK cells in MDD (3). These increased cell numbers could reflect compensatory mechanisms due to impaired cell-intrinsic functions. This idea is supported by functional studies which show reduced NK cytotoxicity (43) and impaired lymphocyte proliferative responses (44–46) in MDD.

In female-specific analyses, pathway analyses highlighted decreases in mitochondrial electron transport chain (oxidative phosphorylation) genes. Additionally, antioxidant genes *GPX1* and *GPX4* (also genetically prioritized in MDD) were decreased in female MDD, aligning with prior findings of impaired antioxidant defenses in depression both peripherally and in the brain (47, 48).

We discovered a consistent decrease in replication-dependent histone expression in MDD, including polyadenylated forms, with these results also significant for both males and females in the sex-stratified meta-analyses. Histones are proteins essential for packaging DNA into chromatin, and regulating gene expression and DNA replication. Normally expressed during DNA replication, these histones can also become polyadenylated in mature tissues (49) and following DNA damage (50, 51). Given the established link between MDD and DNA damage (52), our results may reflect such DNA damage-induced decreases in histones, underscoring histone regulation as an underexplored area in MDD biology.

Consensus network analysis identified multiple differentially expressed gene modules, notably a module featuring the splicing factor *PUF60*, upregulated consistently in MDD in both sexes. As *PUF60* negatively regulates pro-inflammatory cytokine expression (53) this increase may represent a compensatory response to the chronic inflammation often observed in MDD.

### Subtle expression signature and partial concordance with genetically-predicted effects

Our mega-analysis (N=6,000) is large for this data type. However, with the inclusion of mechanisms to ensure robustness - i.e. harmonized reprocessing, correction for cellular composition, and correction for bias and inflation - we find few genes reproducibly associated with depression as a diagnosis. This contrasts with smaller studies reporting extensive differential expression (38, 54). This limited reproducibility likely stems from heterogeneity in depression diagnoses and patient populations, and varying influences of confounding factors, including antidepressant use and symptom severity. Future mega-analyses focusing on biologically or clinically defined subgroups, such as those with inflammatory profiles (55, 56), might yield clearer signatures. Additionally, the underrepresentation of RNA-seq datasets limited transcript-level analyses. Increased availability of RNA-seq data will be crucial, as highlighted by our finding of MDD-associated changes in translation-initiation pathways observable only at the transcript-level.

At a cellular level, our results converged well with genetic predictions, as discussed above. However, at the individual gene level, concordance between our transcriptomic results and genetically predicted changes (via TWAS) was minimal. This discrepancy might reflect limitations of current TWAS methodologies (57), reflect earlier developmental effects of risk genes, or indicate that most MDD-associated transcriptomic alterations result from environmental influences or downstream regulatory changes rather than direct genetic risk.

### Limitations and future directions

Most participants in our analysis were of European ancestry, limiting generalizability. Furthermore, results were mainly driven by the largest dataset, dbGaP. Male-specific analyses lacked statistical power compared to females, emphasizing the necessity for adequately powered sex-stratified studies. Variations in smaller immune cell subsets, which bioinformatics cannot fully adjust for, may have influenced our results (14). Future studies using sorted-cell or single-cell approaches will be essential.

Our results underscore the value of large-scale, harmonized transcriptomic analyses in clarifying the biological underpinnings of depression. While gene-level associations remain modest, converging evidence implicates immune cell dysfunction, mitochondrial stress, and altered chromatin regulation as key processes in MDD. These findings suggest that peripheral gene expression can capture relevant aspects of depression biology. Future studies integrating transcriptomic, proteomic, and functional data may offer new insights into pathophysiology and pave the way for more targeted interventions.

## Data and code availability

Code to reproduce the individual dataset analyses, meta-analyses and figures is available at https://github.com/lynalllab/depression_transcriptome. At the Zenodo repository https://doi.org/10.5281/zenodo.15290507, we provide genome-wide gene- and transcript-level summary statistics; where permitted, we also supply harmonized processed individual-level count matrices along with estimated cell counts and metadata (where permitted).

## Data Availability

Code to reproduce the individual dataset analyses, meta-analyses and figures is available at https://github.com/lynalllab/depression_transcriptome. At the Zenodo repository https://doi.org/10.5281/zenodo.15290507, we provide genome-wide gene- and transcript-level summary statistics; where permitted, we also supply harmonized processed individual-level count matrices along with estimated cell counts and metadata (where permitted).

https://github.com/lynalllab/depression_transcriptome

## Acknowledgement

CE was supported by a Dr Manmohan Singh scholarship, St John’s College, University of Cambridge. MEL was supported by an NIHR Clinical Lectureship. This work was also supported by the ImmunoMIND hub (MR/Z50354X/1) as part of the UKRI Mental Health Platform. All research at the Department of Psychiatry in the University of Cambridge is supported by the NIHR Cambridge Biomedical Research Centre (NIHR203312) and the NIHR Applied Research Collaboration East of England. The views expressed are those of the author(s) and not necessarily those of the NIHR or the Department of Health and Social Care. The dataset(s) used for the analyses described in this manuscript were obtained from the database of Genotypes and Phenotypes (dbGaP) found at http://www.ncbi.nlm.nih.gov/gap through dbGaP accession number phs000486.v1.p1. Samples and associated phenotype data for the GAIN Major Depression: Stage 1 Genome-wide Association In Population Based Samples Study (PI: Dr. Patrick F. Sullivan, MD, University of North Carolina) were provided by Dr. Dorret I. Boomsma, PhD and Dr. Eco de Geus, PhD VU University Amsterdam (PIs NTR), Dr. Brenda W. Penninx, PhD, VU University Medical Center Amsterdam, Dr. Frans Zitman, MD PhD, Leiden University Medical Center, Leiden, and Dr. Willem Nolen, MD PhD, University Medical Center Groningen (PIs and site-PIs NESDA). Funding support for the GAIN Major Depression: Stage 1 Genome-wide Association In Population Based Samples Study (parent studies: Netherlands Study of Depression and Anxiety (NESDA) and the Netherlands Twin Register (NTR)) was provided by the Netherlands Scientific Organization (904-61-090, 904-61-193, 480-04-004, 400-05-717, NWO Genomics, SPI 56-464-1419) the Centre for Neurogenomics and Cognitive Research (CNCR-VU); the European Union (EU/WLRT-2001-01254), ZonMW (geestkracht program, 10-000-1002), NIMH (RO1 MH059160) and matching funds from participating institutes in NESDA and NTR, and the genotyping of samples was provided through the Genetic Association Information Network (GAIN). The authors would like to thank Petra Vertes, Gayle Wittenberg, Jon Greene, Brett McKinney, Georgina Navoly and Karoline Kuchenbaecker for helpful discussions and assistance in the preparation of this work.

