## Supplementary Material for "Systematic review and mega-analysis of the peripheral blood transcriptome in depression implicates dysregulation of lymphoid cells and histones"

### Supplementary Figures

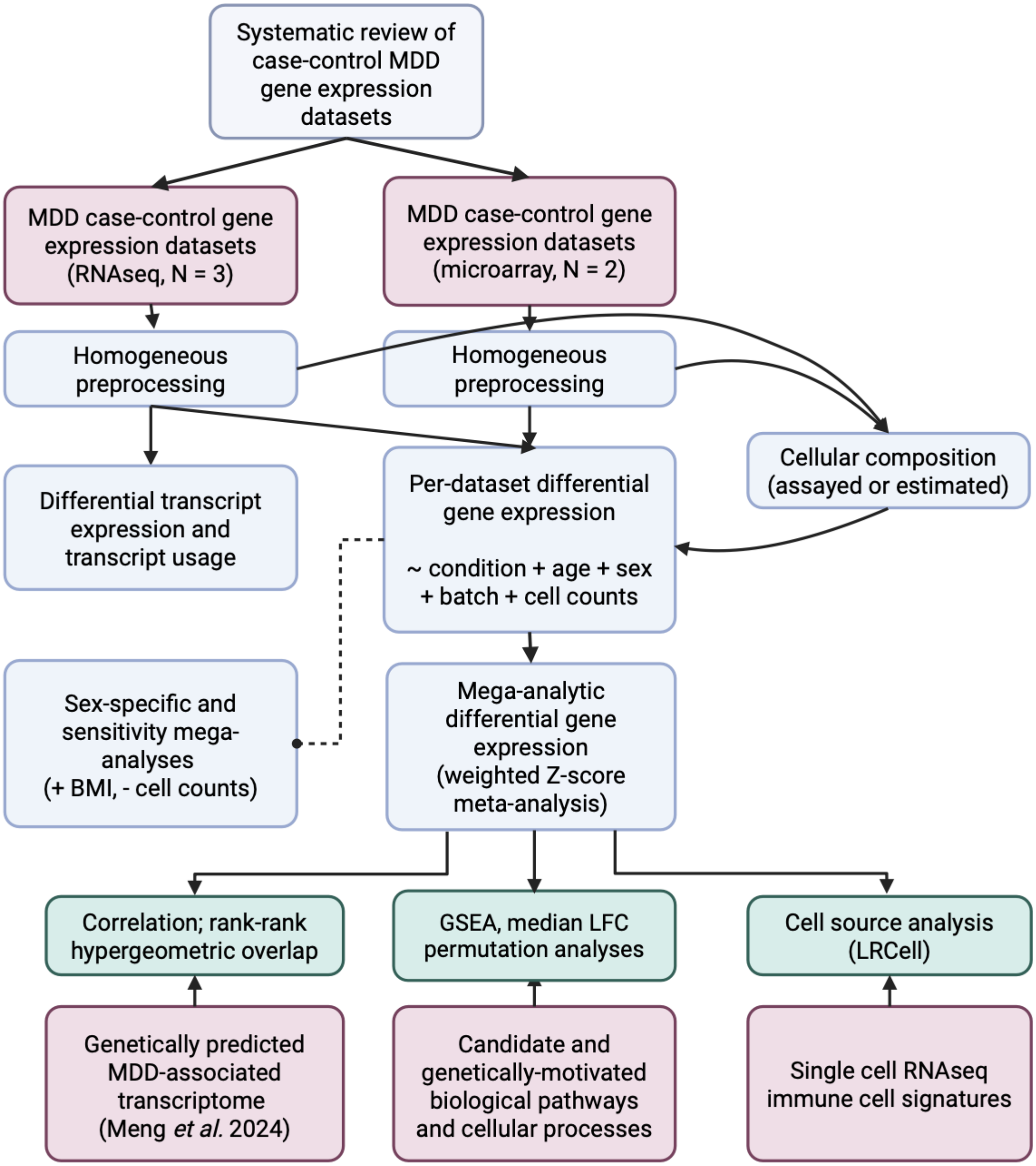


***Figure S1. Overview of main analytic steps for transcriptome mega-analysis.*** *See **Figure S2** for in depth schematic of expression data analysis and **Figure S3** for further details of systematic review (PRISMA diagram). GSEA, gene set enrichment analysis; LFC, log fold change; MDD, major depressive disorder.*

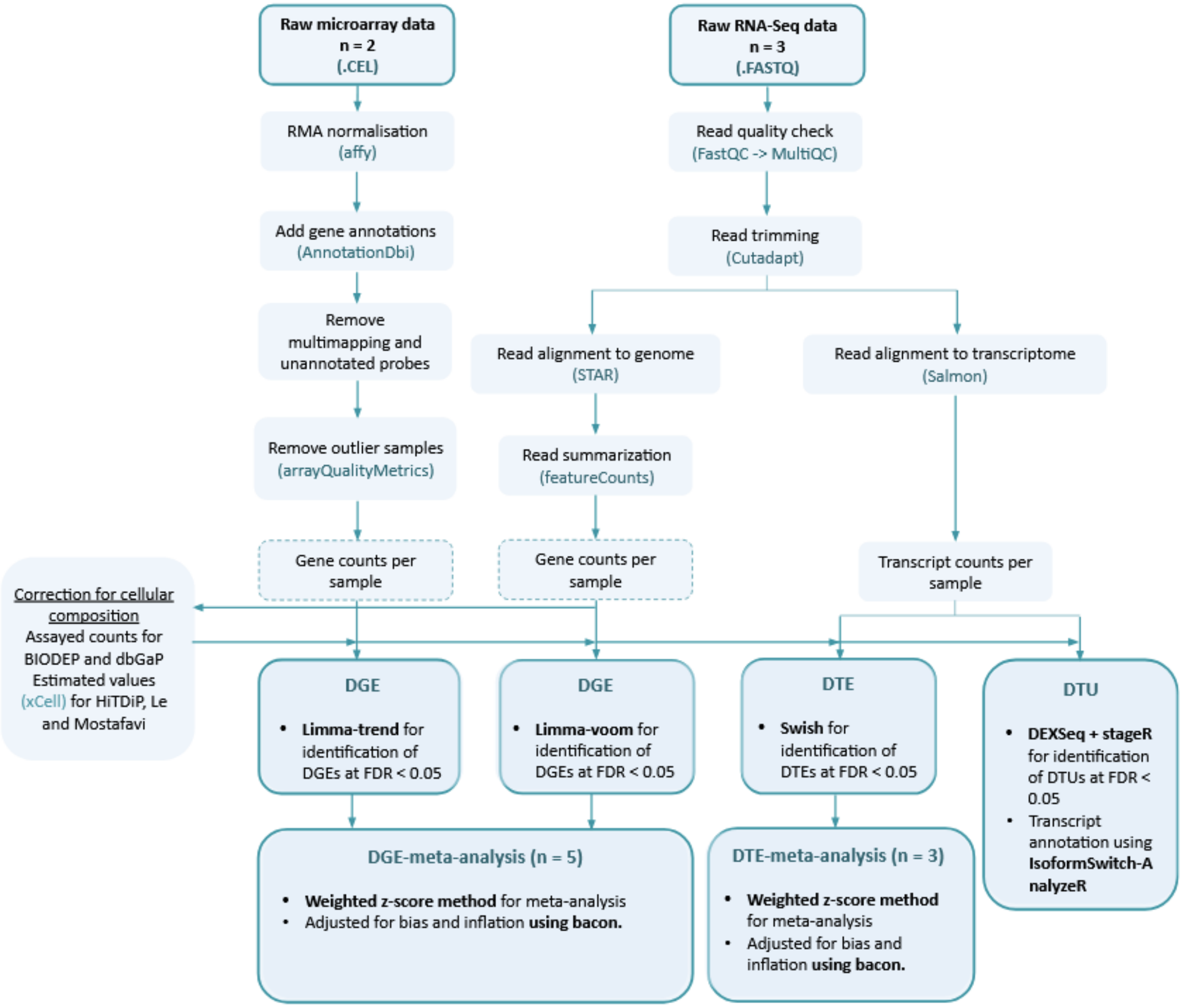


***Figure S2 Pipeline for processing gene and transcript expression data obtained from raw RNA-Seq or microarray technologies***

*A standardized analytical pipeline was used to process the five MDD case-control datasets. Diagram shows main pipeline steps for each data type, and the file formats and software tools used. For DGE, RNA-Seq and microarray were initially processed separately for gene quantification. Gene expression was modelled for each dataset separately, then results were meta-analyzed, including both RNA-seq and microarray datasets in the meta-analysis. For DTE and DTU, only RNA-seq datasets can be analyzed. RMA, Robust Multi-array Average method; STAR, Spliced Transcripts Alignment to a Reference; DGE, Differential gene expression; DTE, Differential transcript expression; DTU, Differential transcript usage; FDR, False discovery rate.*

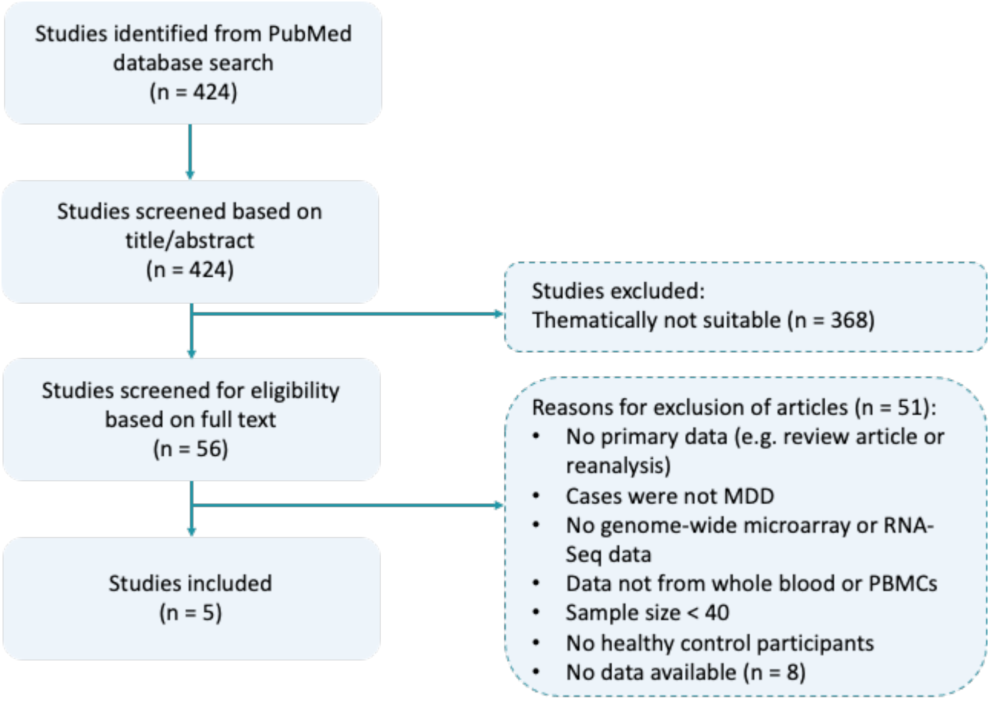


***Figure S3. PRISMA diagram for study selection***

*Number of studies screened and excluded at each stage of the review process have been highlighted. Based on a search of 424 research articles from the PubMed database, five MDD case-control datasets were identified for meta-analysis.*

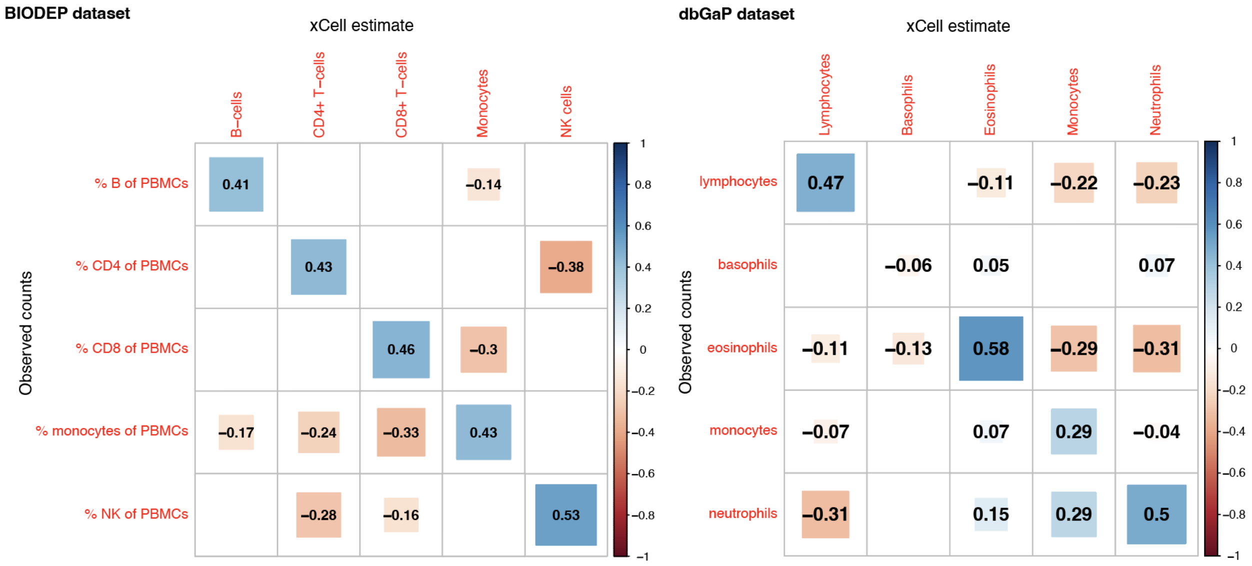


***Figure S4. Correlation between computationally generated cell type enrichment scores and assayed cell count values for PBMC and whole blood MDD case-control datasets***

*Differences in cell count proportions can influence gene expression values, however, cell count information was available for only a subset of the five MDD case-control datasets. Cellular deconvolution (using xCell, see **Methods**) was used to infer cell type enrichment from gene expression data. To validate these estimates, Spearman's correlation was estimated between original and xCell cell count values for the subsets of dataset with cell count information: the BIODEP PBMC MDD case-control dataset (left) and the subset of the dbGaP case-control whole blood dataset for which cell counts were available (right). Tile color indicates strength of correlation (rho), see scale bar; tile size indicates magnitude of rho. Tiles with non-significant p-values (P >= 0.05) are blank. Sample sizes: BIODEP: 211 samples; dbGaP: 2,868 samples.*

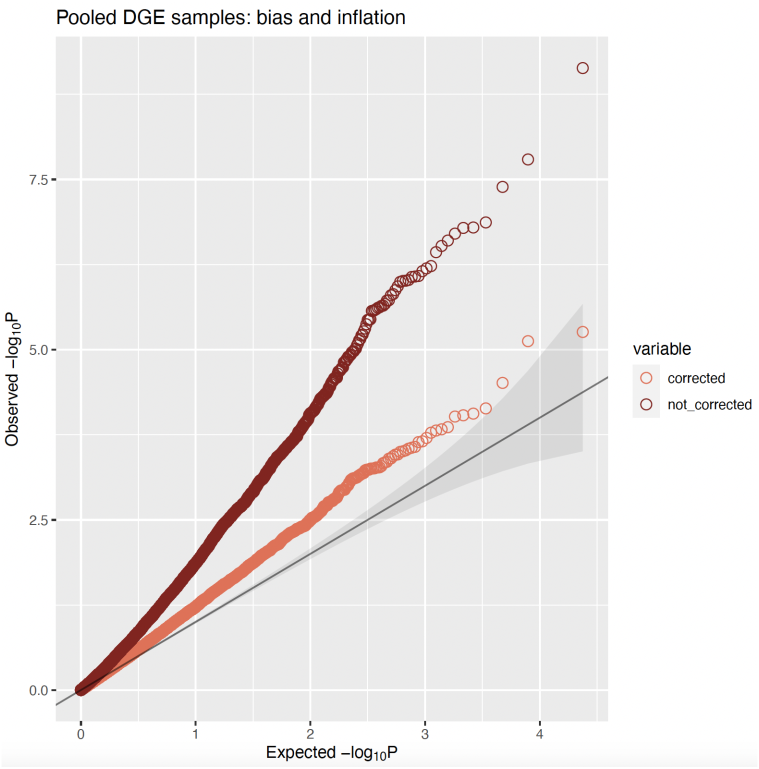


***Figure S5. QQ plot for pooled analysis differential gene expression***

*QQ plot for bias- and inflation-corrected pooled DGE results with and without correction for cellular composition. Dark red dots correspond to DGE meta-analytic results where per-dataset DGE models did not correct for cellular composition; coral dots correspond to meta-analytic results where per-dataset models did correct for cellular composition. Black line indicates the expected (uniform) distribution of p-values under the null hypothesis.*

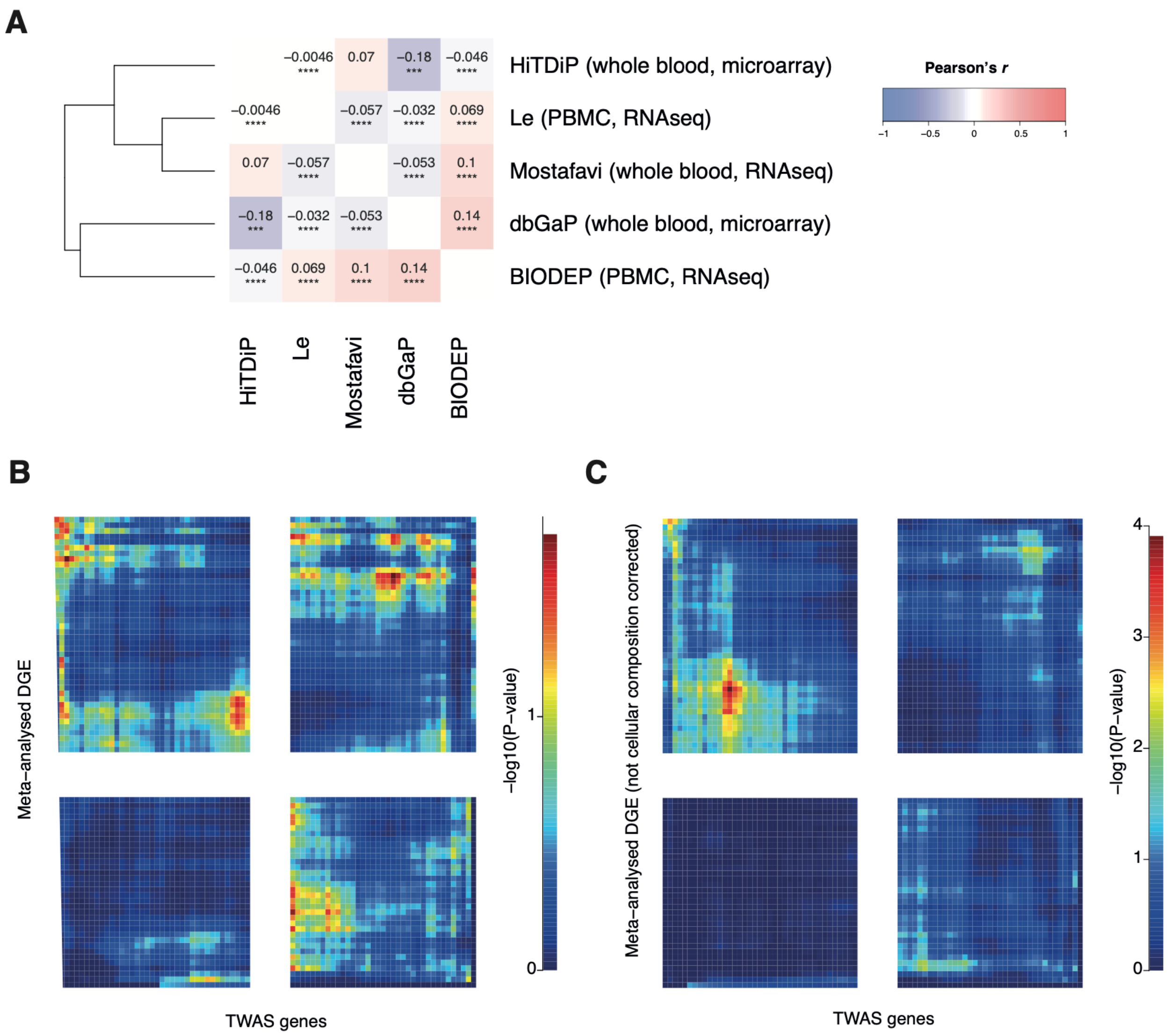


***Figure S6. Concordance of the pattern of gene expression results across datasets and with genetically predicted results*** *(A) Correlation plot shows bivariate correlations between individual dataset DGE results; numbers on tiles show Pearson's r; dendrogram shows hierarchical clustering of datasets based on 'Euclidean' distance and the 'complete' hclust method; significance is shown as **** P< 0.0001; *** P >= 0.0001 & P<0.001; ** P>= 0.001 & P < 0.01; * P>= 0.01 & P < 0.05; (B) DGE concordance between TWAS and cell corrected MDD meta-analysis shown using RRHO plot (see* ***Methods****); Pearson correlation between observed DGE and genetically predicted DGE is r=0.0015, p=0.91 (same as* ***Figure 3D****, shown again for comparison). (C) DGE concordance between TWAS and non-cell corrected MDD meta-analysis using RRHO plot; Pearson correlation between observed DGE and genetically predicted DGE is r=0.0024, p=0.86*

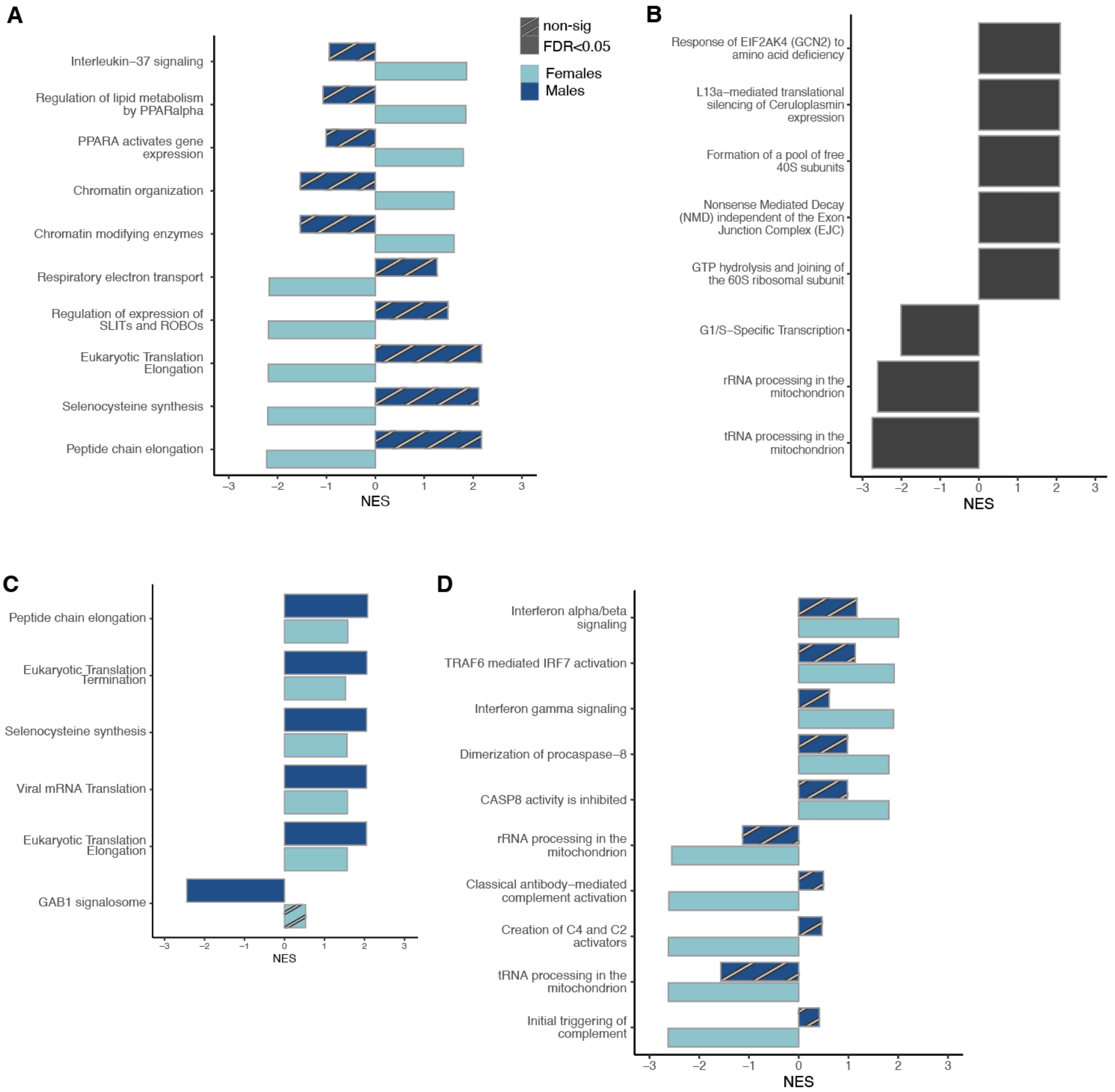


***Figure S7. Additional meta-analytic Reactome pathway enrichment results*** *Graphs show the 5 most upregulated pathways (by NES scores) and 5 most downregulated pathways (by NES score) for each meta-analysis. Only pathways where FDR<0.05 are shown, so some plots show fewer than 5 downregulated pathways. (A) female-only DGE meta-analysis (no Reactome pathways were enriched at FDR<0.05 in the male-only DGE meta-analysis) (B) pooled-sex DTE meta-analysis (C) male-only DTE meta-analysis (D) female-only DTE meta-analysis. For (B,C,D), results are based on the datasets for which transcript-level information was available i.e. RNA sequencing datasets (N=691 MDD, N=569 controls; from BIODEP, Le and Mostafavi datasets).*

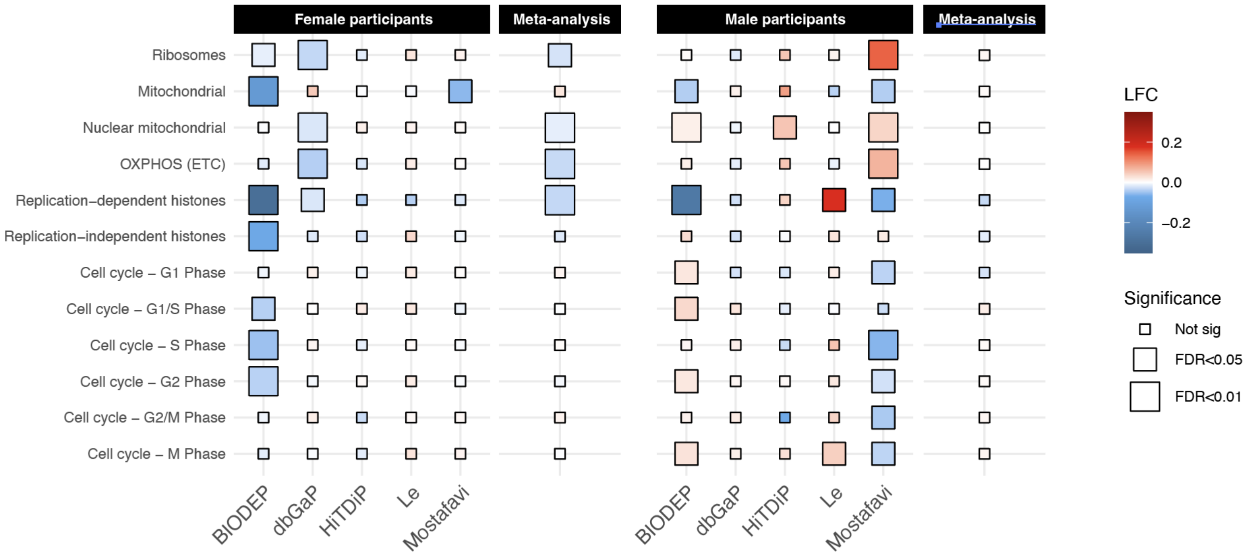


***Figure S8. Sex-stratified permutation analysis of core cellular processes*** *Heatmaps show MDD-associated changes in median expression of genes involved in core cellular processes for female and male samples. A permutation test was conducted (where genes with similar expression was used as replacements, see* ***Supplementary Text****) to calculate p-values. Size of the rectangles indicates these adjusted BH-corrected p-values (FDR). For each dataset, tile color indicates median LFC for genes present in a specific gene group and meta-analytic tile color shows the mean of these LFCs across datasets, weighted by sample size. Per-dataset permutation values were meta-analyzed using the weighted Z-score method. ETC, electron transport chain; G, Gap or growth phase; S, Synthesis phase; M, Mitotic phase.*

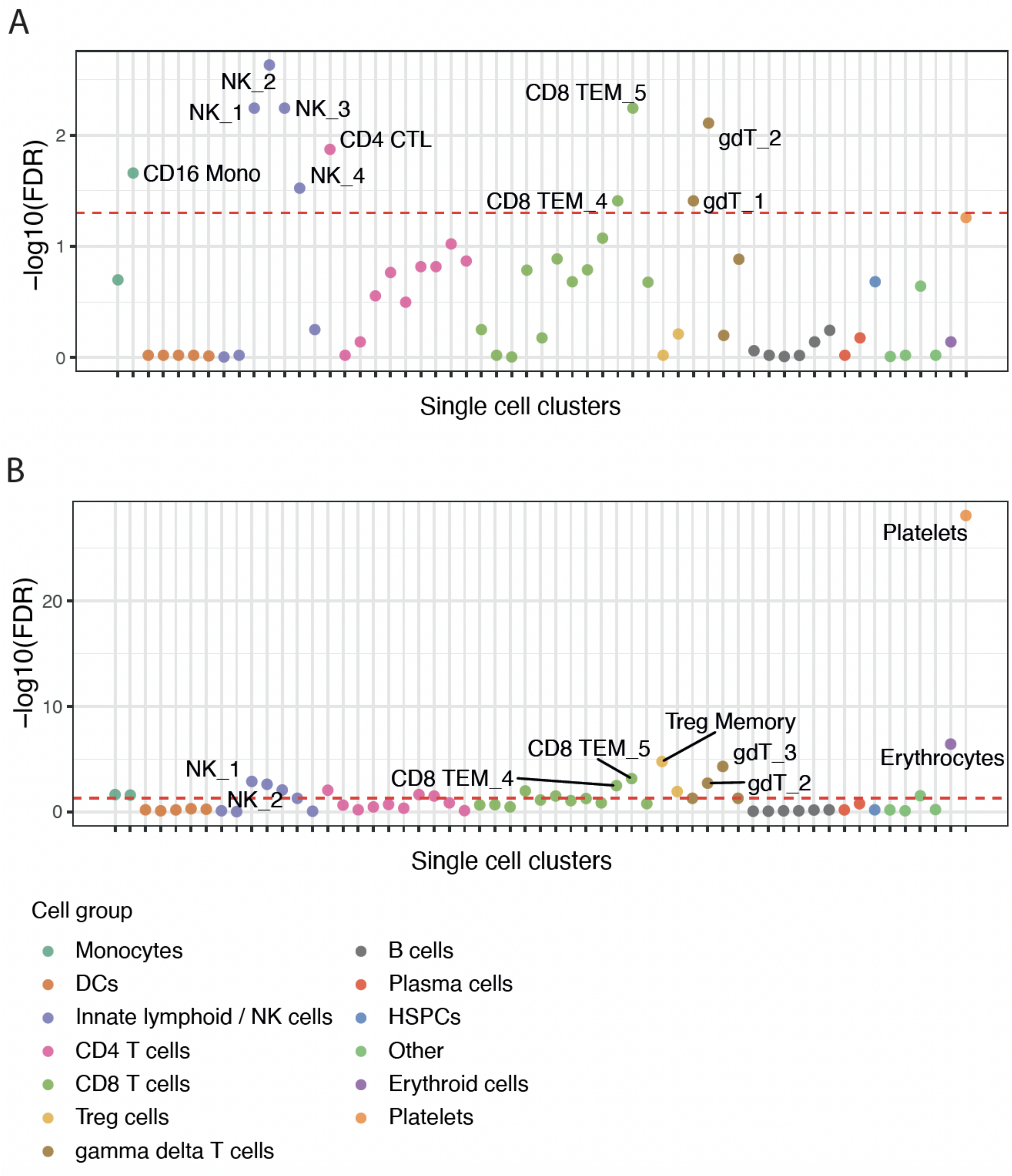


***Figure S9. Cell origin analysis with LRCell*** *Cell origin analysis indicates the cell subsets estimated to be driving the MDD-associated expression signature. Dot plot shows regression of the MDD expression signature (sex-pooled analysis) against cell subset marker genes derived from a peripheral blood single cell dataset. X-axis shows the immune cell types tested, with legend showing the major cell subgroups. Y-axis shows the significance of each cell type enrichment by -log10(FDR); dashed red line indicates FDR=0.05; the most significant enrichments are labelled. (A) Cell origin analysis additionally including estimated platelet counts in whole-blood models (B) Cell origin analysis additionally including BMI in DGE models.*

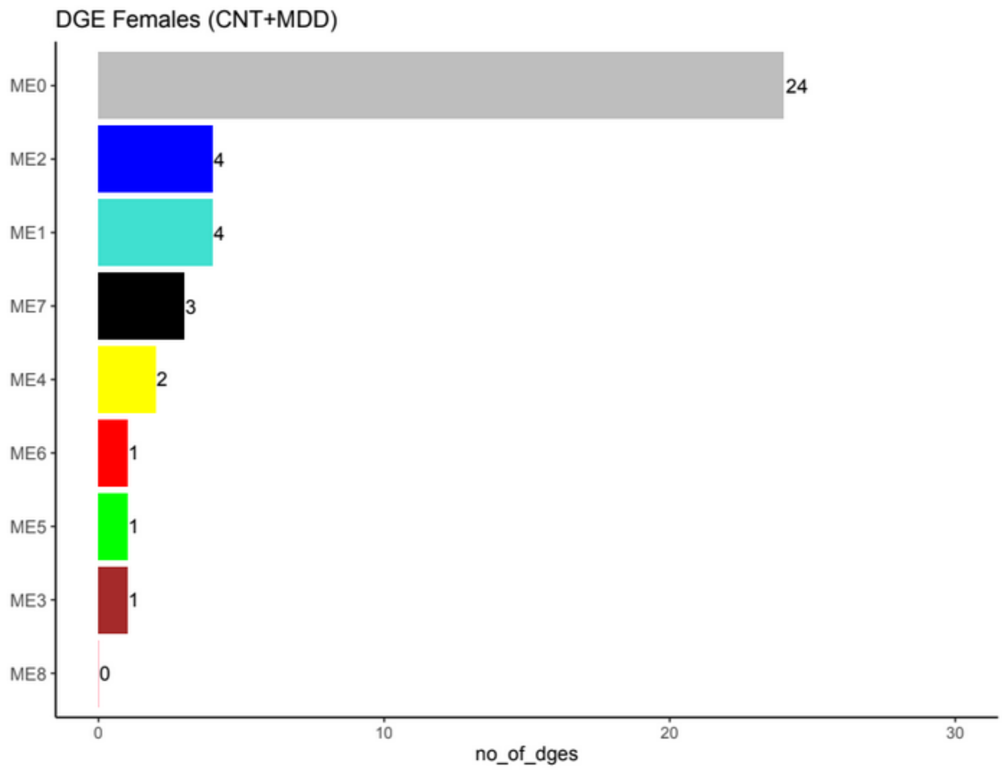


***Figure S10 Module membership of meta-analytic DGE genes.*** *Significant DGE and DTE results from the female DGE analysis were evaluated for membership in the female WGCNA consensus modules generated (there were no genome-wide significant DGE or DTE results in males). Module names are shown on the y-axes and the number of differentially expressed genes (x-axes) are shown next to the bar plots. Grey module indicates genes not assigned to any module.*

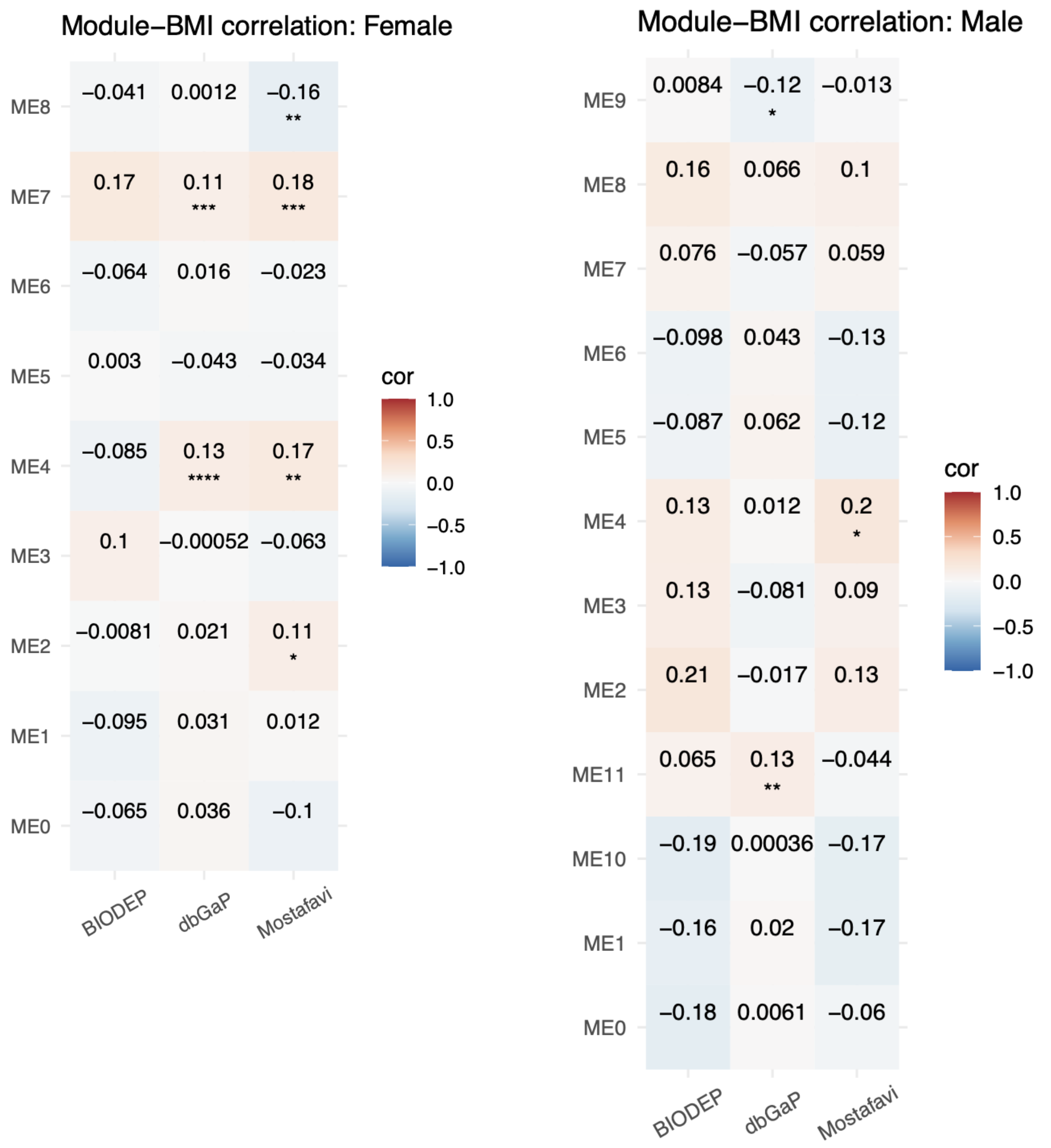


***Figure S11. Association between male and female MDD case-control consensus modules and BMI***

*Pearson correlations were used to evaluate association between consensus module eigengenes (x-axes) and BMI in datasets where BMI was ascertained (y-axes). Tile color shows Pearson's r, with blue indicating positive correlations and red indicating negative correlations. Significance is indicated as **** P< 0.0001; *** P >= 0.0001 & P<0.001; ** P>= 0.001 & P < 0.01; * P>= 0.01 & P < 0.05. Sample sizes used in this analysis are as follows: CNT + MDD females: BIODEP = 143 samples, Mostafavi = 622 samples, dbGaP = 2,983 samples, and CNT + MDD males: BIODEP = 62 samples, Mostafavi = 263 samples, dbGaP = 1,532 samples.*

### Supplementary Tables

***Table S1. Covariate information for the five MDD case-control datasets and association of covariates with disorder status*** *Data are shown for the five MDD case-control datasets included in the mega-analysis, with total sample number for cases (MDD) and controls (CNT) in brackets. Covariate values are shown for categorical variables as total number (and as a percentage of total sample group in brackets), and shown for continuous variables as mean values (with standard deviation in brackets). P-values for association of each covariate with MDD status were evaluated using a t-test for continuous covariates and a Chi-squared test for categorical covariates. '-' denotes missing data.*

<table>
<tr><th rowspan="2">Covariates</th><th colspan="3">BIODEP</th><th colspan="3">dbGaP</th><th colspan="3">HiTDiP</th><th colspan="3">Le</th><th colspan="3">Mostafavi</th></tr>
<tr><th>CNT (N=47)</th><th>MDD (N=164)</th><th>P</th><th>CNT (N=2976)</th><th>MDD (N=1564)</th><th>P</th><th>CNT (N=61)</th><th>MDD (N=120)</th><th>P</th><th>CNT (N=78)</th><th>MDD (N=75)</th><th>P</th><th>CNT (N=444)</th><th>MDD (N=452)</th><th>P</th></tr>
<tr><td>Sex: F</td><td>32 (68.1%)</td><td>114 (69.5%)</td><td rowspan="2">0.994</td><td>1918 (64.4%)</td><td>1083 (69.2%)</td><td rowspan="2">0.00132</td><td>45 (73.8%)</td><td>89 (74.2%)</td><td rowspan="2">1</td><td>39 (50.0%)</td><td>50 (66.7%)</td><td rowspan="2">0.0542</td><td>277 (62.4%)</td><td>353 (78.1%)</td><td rowspan="2"><0.001</td></tr>
<tr><td>Sex: M</td><td>15 (31.9%)</td><td>50 (30.5%)</td><td>1058 (35.6%)</td><td>481 (30.8%)</td><td>16 (26.2%)</td><td>31 (25.8%)</td><td>39 (50.0%)</td><td>25 (33.3%)</td><td>167 (37.6%)</td><td>99 (21.9%)</td></tr>
<tr><td>Batch</td><td>9|12|8|8|10</td><td>26|55|19|33|31</td><td>0.716</td><td>1918|1058</td><td>1083|481</td><td>0.00132</td><td>29|32</td><td>57|63</td><td>1</td><td>20|21|17|20</td><td>18|18|22|17</td><td>0.762</td><td>220|224</td><td>240|212</td><td>0.32</td></tr>
<tr><td>Age</td><td>33.90 (±6.72)</td><td>35.89 (±7.94)</td><td>0.0897</td><td>35.87 (±2.32)</td><td>41.62 (±12.41)</td><td><0.001</td><td>52.01 (±11.54)</td><td>52.57 (±11.52)</td><td>0.759</td><td>30.74 (±10.47)</td><td>33.04 (±10.62)</td><td>0.18</td><td>44.55 (±11.02)</td><td>44.84 (±10.64)</td><td>0.694</td></tr>
<tr><td>BMI</td><td>25.41 (±4.87)</td><td>27.12 (±6.21)</td><td>0.0538</td><td>24.18 (±4.34)</td><td>25.72 (±5.27)</td><td><0.001</td><td>-</td><td>-</td><td>-</td><td>-</td><td>-</td><td>-</td><td>27.93 (±6.51)</td><td>30.20 (±7.71)</td><td><0.001</td></tr>
<tr><td>Time blood drawn</td><td>-</td><td>-</td><td>-</td><td>8.5 (±1.2)</td><td>8.6 (±1.32)</td><td>0.00248</td><td>-</td><td>-</td><td>-</td><td>-</td><td>-</td><td>-</td><td>12.7 (±3.53)</td><td>12.44 (±3.31)</td><td>0.264</td></tr>
<tr><td>Smoking: Yes</td><td>-</td><td>-</td><td>-</td><td>672 (22.6%)</td><td>674 (43.1%)</td><td rowspan="2"><0.001</td><td>-</td><td>-</td><td>-</td><td>-</td><td>-</td><td>-</td><td>34 (7.7%)</td><td>80 (17.7%)</td><td rowspan="2"><0.001</td></tr>
<tr><td>Smoking: No</td><td>-</td><td>-</td><td>-</td><td>2303 (77.4%)</td><td>890 (56.9%)</td><td>-</td><td>-</td><td>-</td><td>-</td><td>-</td><td>-</td><td>410 (92.3%)</td><td>372 (82.3%)</td></tr>
</table>

***Table S2. Cell counts in depression and controls in each dataset.*** *All values are xCell-estimated proportions, except values with an asterisk, which were directly assayed. Mean values (with standard deviation in brackets) for MDD cases and controls are shown for the five datasets. Cell count values were not available for 30 BIODEP samples. P-values for association of each covariate with MDD were evaluated using a t-test. '-' indicates the data type is not applicable (i.e. the cell type is not expected to be present in the sample).*

| Covariates | BIODEP | | | dbGaP | | | HiTDiP | | | Le | | | Mostafavi | | |
|---|---|---|---|---|---|---|---|---|---|---|---|---|---|---|---|
| | CNT (N=47) | MDD (N=164) | P | CNT (N=2976) | MDD (N=1564) | P | CNT (N=61) | MDD (N=120) | P | CNT (N=78) | MDD (N=75) | P | CNT (N=444) | MDD (N=452) | P |
| B-cells | *4.14 (±2.92) | *4.68 (±3.45) | 0.292 | 0.0698 (±0.0738) | 0.0694 (±0.0732) | 0.857 | 0.18 (±0.0882) | 0.14 (±0.0821) | 0.011 | 0.08 (±0.07) | 0.10 (±0.06) | 0.0159 | 0.307 (±0.115) | 0.312 (±0.121) | 0.551 |
| CD4+ T-cells | *46.56 (±10.70) | *47.31 (±11.09) | 0.678 | 0.33 (±0.08) | 0.35 (±0.07) | <0.001 | 0.21 (±0.0771) | 0.19 (±0.0774) | 0.18 | 0.0222 (±0.03) | 0.0169 (±0.02) | 0.177 | 0.10 (±0.05) | 0.11 (±0.06) | 0.167 |
| CD8+ T-cells | *20.78 (±4.82) | *19.04 (±7.04) | 0.0539 | 0.32 (±0.07) | 0.29 (±0.08) | <0.001 | 0.19 (±0.05) | 0.15 (±0.07) | <0.001 | 0.04 (±0.0312) | 0.05 (±0.0334) | 0.174 | 0.1471 (±0.059) | 0.1497 (±0.055) | 0.5 |
| NK cells | *5.60 (±3.43) | *6.83 (±4.93) | 0.0528 | 0.25 (±0.106) | 0.23 (±0.107) | <0.001 | 0.16 (±0.07052) | 0.13 (±0.07095) | 0.032 | 0.055 (±0.05) | 0.048 (±0.04) | 0.285 | 0.0643 (±0.0427) | 0.0586 (± 0.0402) | 0.04 |
| Monocytes | *12.30 (±6.32) | *13.34 (±6.14) | 0.32 | 0.03 (±0.0404) | 0.04 (±0.0437) | <0.001 | 0.0008 (±0.003) | 0.004 (±0.01) | 0.0172 | 0.0319 (±0.0238) | 0.0316 (±0.0225) | 0.935 | 0.006 (±0.0086) | 0.005 (±0.0081) | 0.538 |
| Eosinophils | - | - | - | 0.76 (±0.24) | 0.734 (±0.22) | <0.001 | 0.69 (±0.21) | 0.76 (±0.28) | 0.0613 | - | - | - | 0.1130 (±0.062) | 0.105 (±0.064) | 0.07 |
| Neutrophils | - | - | - | 0.21 (±0.04) | 0.22 (±0.03) | <0.001 | 0.07 (±0.054) | 0.08 (±0.0496) | 0.305 | - | - | - | 0.0002 (±0.001) | 0.0003 (±0.002) | 0.36 |
| Platelets | - | - | - | 0.133 (±0.0472) | 0.125 (±0.0455) | <0.001 | 0.14 (±0.066) | 0.16 (±0.072) | 0.017 | - | - | - | 0.148 (±0.0889) | 0.146 (±0.0872) | 0.749 |

***Table S3. Gene co-expression modules identified using WGCNA*** *Modules do not include genes that could not be assigned to a module (grey on WGCNA dendrograms).*

| Dendrogram type | Number of samples used | Number of modules | Minimum module size | Maximum module size | Average module size |
|---|---|---|---|---|---|
| **Females** | 3967 | 8 | 75 | 864 | 332 |
| **Males** | 1966 | 11 | 127 | 641 | 314 |

***Table S4. Similarity of WGCNA modules across datasets*** *Table shows mean preservation scores between each pair of datasets (see* ***Supplementary Text****). These scores representing the similarity between the same inter-dataset module (e.g. between eigengenes for ME1 in BIODEP and ME1 in Le dataset), compared to the similarity to all other consensus module eigengenes (e.g. between eigengenes for ME1 module in BIODEP and all other (non-ME1) module eigengenes in the Le dataset). Values closer to one indicate greater inter-dataset similarity.*

| **Females** | **BIODEP** | **Le** | **Mostafavi** | **dbGaP** | **HiTDiP** |
|---|---|---|---|---|---|
| **BIODEP** | 1 | 0.87 | 0.89 | 0.93 | 0.93 |
| **Le** | 0.87 | 1 | 0.83 | 0.83 | 0.84 |
| **Mostafavi** | 0.89 | 0.83 | 1 | 0.87 | 0.87 |
| **dbGaP** | 0.93 | 0.83 | 0.87 | 1 | 0.96 |
| **HiTDiP** | 0.93 | 0.84 | 0.87 | 0.96 | 1 |

| **Males** | **BIODEP** | **Le** | **Mostafavi** | **dbGaP** | **HiTDiP** |
|---|---|---|---|---|---|
| **BIODEP** | 1 | 0.85 | 0.87 | 0.88 | 0.91 |
| **Le** | 0.85 | 1 | 0.83 | 0.82 | 0.81 |
| **Mostafavi** | 0.87 | 0.83 | 1 | 0.84 | 0.85 |
| **dbGaP** | 0.88 | 0.82 | 0.84 | 1 | 0.93 |
| **HiTDiP** | 0.91 | 0.81 | 0.85 | 0.93 | 1 |

***Table S5. MDD-associated co-expression modules in WGCNA consensus networks in males (top) and females (below)*** *Table shows results of differential module expression meta-analysis, comparing the value of per-module eigengenes between MDD cases and controls using a t test, followed by weighted Z-score meta-analysis (see* ***Methods****). Only modules showing significant meta-analytic association with MDD status at FDR<0.05 are shown. Effect sizes are indicated by the average weighted Cohen's d. The top hub genes i.e. genes with the highest connectivity (weighted module membership, kME, weighted by sample size of the individual dataset), are also shown. Female analysis = 3,967 samples; Male analysis = 1,966 samples.*

| **Male consensus modules** | **Top module pathway** | **Average weighted Cohen's d** | **Meta-analytic p-value** | **Module hub gene** | **Gene Description** | **Potential Hub gene function** | **Weighted kME** |
|---|---|---|---|---|---|---|---|
| **ME9** | Myeloid cell activation involved in immune response | 0.152 | 1.5e-03 | *RPS6KA1* | Ribosomal protein S6 Kinase A1 | Kinase involved in cell growth and differentiation | 0.77 |
| **ME11** | Response to wounding | 0.1 | 1.3e-02 | *CLU* | Clusterin | Extracellular molecular chaperone involved in several biological processes such as cell death and tumor progression | 0.83 |
| **ME3** | Myeloid leukocyte mediated immunity | 0.1 | 3.88e-03 | *PUF60* | Poly(U) binding splicing factor 60 | Nuclear processes such as pre-mRNA splicing, apoptosis, and transcriptional regulation | 0.80 |

| Female consensus modules | Top module pathway | Average weighted Cohen's d | Meta-analytic p-value | Module hub gene | Gene Description | Potential Hub gene function | Weighted kME |
|---|---|---|---|---|---|---|---|
| **ME7** | Response to wounding | 0.209 | 3.64e-12 | *GMPR* | Guanosine monophosphate reductase | Deamination of GMP to inosine monophosphate (IMP) | 0.83 |
| **ME4** | Protein localization to membrane | 0.179 | 9.19e-11 | *COX4I1* | Cytochrome C oxidase subunit IV isoform 1 | Electron transfer and proton pumping in mitochondrial respiratory chain | 0.78 |
| **ME2** | SCF-dependent proteasomal ubiquitin-dependent protein catabolic process | 0.160 | 1.5e-08 | *UBXN1* | UBX domain protein 1 | Negative regulation of protein metabolic process and NF-kB signaling | 0.77 |
| **ME5** | Leukocyte degranulation | 0.111 | 1.58e-05 | *PUF60* | Poly(U) binding splicing factor 60 | Nuclear processes such as pre-mRNA splicing, apoptosis, and transcriptional regulation | 0.77 |
| **ME1** | Endomembrane system organization | -0.105 | 1.11e-03 | *MARCH7* | Membrane associated ring-CH-type finger 7 | Protein degradation | 0.80 |
| **ME6** | mRNA processing | -0.120 | 6.91e-06 | *TTC14* | Tetratricopeptide repeat domain 14 | Potentially involved in nucleic acid binding | 0.75 |

***Table S6. MDD-associated co-expression modules in male and female WGCNA consensus network - Leave One Out (LOO) analysis.*** *Table shows weighted Z-score meta-analysis excluding dbGaP (the largest dataset), showing that differential module expression results (**Figure 3**) were primarily driven by dbGaP; MDD-associated modules were no longer significant in a leave-one-out meta-analysis without this dataset.*

| **Male modules** | **P Value** | **Direction of effect** | **FDR** | **Average weighted Cohen's d** |
|---|---|---|---|---|
| ME1 | 0.11 | + | 0.19 | 0.11 |
| ME2 | 0.004 | - | 0.045 | -0.28 |
| ME3 | 0.87 | - | 0.87 | -0.07 |
| ME4 | 0.02 | - | 0.09 | -0.19 |
| ME5 | 0.41 | + | 0.50 | 0.02 |
| ME6 | 0.14 | + | 0.21 | 0.19 |
| ME7 | 0.36 | + | 0.50 | 0.11 |
| ME8 | 0.05 | - | 0.14 | -0.20 |
| ME9 | 0.06 | + | 0.14 | 0.17 |
| ME10 | 0.07 | + | 0.14 | 0.18 |
| ME11 | 0.83 | + | 0.87 | -0.04 |

| **Female module** | **P Value** | **Direction of effect** | **FDR** | **Average weighted Cohen's d** |
|---|---|---|---|---|
| ME1 | 0.38 | - | 0.92 | -0.08 |
| ME2 | 0.38 | + | 0.92 | 0.05 |
| ME3 | 0.85 | - | 0.92 | -0.007 |
| ME4 | 0.63 | + | 0.92 | 0.02 |
| ME5 | 0.89 | + | 0.92 | -0.03 |
| ME6 | 0.67 | - | 0.92 | 0.02 |
| ME7 | 0.92 | + | 0.92 | 0.02 |
| ME8 | 0.33 | - | 0.92 | -0.09 |

***Table S7. Association (t-test) between consensus WGCNA modules and smoking in males and females*** *T-values indicate direction and magnitude of association with current smoking status (yes vs no), for the datasets where smoking status was ascertained (dbGaP and Mostafavi).*

| **Male module** | **dbGaP P Value** | **dbGaP T value** | **Mostafavi P Value** | **Mostafavi T value** |
|---|---|---|---|---|
| ME1 | 0.17 | -1.37 | 0.08 | 1.82 |
| ME2 | 0.49 | 0.70 | 0.34 | -0.97 |
| ME3 | 0.15 | 1.45 | 0.63 | -0.49 |
| ME4 | 0.49 | 0.69 | 0.06 | -1.92 |
| ME5 | 0.17 | 1.28 | 0.32 | 1.0005 |
| ME6 | 0.39 | 0.85 | 0.35 | 0.95 |
| ME7 | 0.05 | 1.96 | 0.45 | -0.76 |
| ME8 | 0.67 | 0.43 | 0.61 | -0.52 |
| ME9 | 0.07 | 1.78 | 0.76 | 0.31 |
| ME10 | 0.25 | -1.16 | 0.08 | 1.81 |
| ME11 | 1.40e-08 | 5.77 | 0.29 | 1.07 |

| **Female module** | **dbGaP P Value** | **dbGaP T value** | **Mostafavi P Value** | **Mostafavi T value** |
|---|---|---|---|---|
| ME1 | 0.95 | 0.06 | 0.31 | 1.03 |
| ME2 | 0.32 | 0.9998 | 0.58 | -0.56 |
| ME3 | 0.02 | 2.38 | 0.54 | -0.62 |
| ME4 | 0.77 | -0.29 | 0.41 | -0.83 |
| ME5 | 0.12 | 1.56 | 0.60 | -0.52 |
| ME6 | 0.43 | 0.78 | 0.32 | 0.998 |
| ME7 | 1.92e-05 | 4.30 | 0.07 | 1.81 |
| ME8 | 0.12 | -1.56 | 0.44 | 0.77 |

***Table S8 DGE mega-analysis additionally corrected for BMI*** *These genome-wide summary statistics are presented as an additional file, showing the BMI-adjusted DGE meta-analysis results for the three MDD case-control datasets where BMI was also ascertained: BIODEP, dbGaP and Mostafavi. Gene expression models were as per the main model (i.e. including cell counts) and additionally included BMI (see* ***Methods****). Column names are as follows: BacWeightedZ_meta indicates bias and inflation corrected and meta-analysed Z-score; pvalue.pval_BacWeightedZ indicates p-value or significance of meta-analysed Z-score; pvalue.BacWeightedZ_adj_pval indicates BH-corrected p-value; EmpiricalEstimatedMean indicates bias calculated using the [R] package bacon; EmpiricalEstimatedSD indicates inflation calculated using the R package bacon.*

### Supplementary Text

#### MDD case-control study selection

A literature review was conducted following the Preferred Reporting Items for Systematic Reviews and Meta-Analyses (PRISMA) guidelines. A comprehensive search of PubMed using the following MeSH terms was performed:

(("major depressive disorder"[Title/Abstract]) OR (MDD[Title/Abstract])) AND ((blood[Title/Abstract]) OR (PBMCs[Title/Abstract]) OR (WBs[Title/Abstract])) AND ((RNA-Seq[Title/Abstract]) OR (RNA-seq[Title/Abstract]) OR (RNA[Title/Abstract]) OR (microarray*[Title/Abstract]) OR (gene*[Title/Abstract]))

The primary inclusion criterion was RNA-Seq or microarray gene expression measurements from blood assayed as part of MDD case-control studies in the adult population. Studies published until August 2021 were considered for inclusion and study selection was performed by two reviewers independently (C.E. and M.E.L.) using the covidence.org platform.

An initial search of the PubMed database (see Methods for search terms) yielded 424 results. Screening of titles and abstracts of studies identified 368 studies as thematically unsuitable as the focus was not on gene expression or MDD. The remaining 56 studies were further assessed for eligibility by full-text review against the inclusion and exclusion criteria provided, identifying 5 primary research articles which met criteria (see PRISMA diagram, **Figure S3** and **Supplementary Text**).

<u>Inclusion criteria:</u>

- Population:
    - >= 18 years
    - Not pregnant
    - Cohorts of healthy controls and MDD cases (for example, not a study of MDD in Alzheimer's patient cohort)
- Concept:
    - Disease outcome was Major Depressive Disorder (MDD)

    - Study evaluates gene or transcript expression changes
    - Gene and transcript expressions were generated using RNA-Seq or Microarrays (mRNA only or genome-wide)
- Context:
    - Any geographical location
    - Language of publication was English
    - Published before 1st August 2021
    - Peer-reviewed research articles
    - Primary studies
    - Blood-based human samples with a minimum of 40 MDD samples

**Sample information**

Each dataset included a variable number of associated metadata comprising continuous variables such as age, body mass index (BMI), and depressive symptom assessment scores, and categorical data on sex, medication history, and presence of comorbid symptoms. Participants with psychosis, schizophrenia, bipolar disorder, history of alcohol or drug abuse, and other major medical disorders were excluded during recruitment. However, each dataset contains a composite of drug naive, treatment-resistant, remitted MDD cases and MDD patients on different medications. One of the studies identified from literature included two different MDD case-control datasets. However, one of the two datasets was not available when directly requested, and therefore, was not included in our analysis. Details of the five datasets included are as below:

1) **BIODEP dataset**: The Biomarkers in Depression (BIODEP) study was conducted as part of Wellcome Trust's Neuroimmunology of Mood Disorders and Alzheimer's disease (NIMA) consortium and consists of control and MDD participants (meeting DSM-V criteria) recruited from five different centers within the United Kingdom. MDD severity was ascertained using the 17-item Hamilton Rating Scale for Depression (HAM-D). Relevant metadata available for this in-house dataset include completed diagnostic and symptom questionnaires, peripheral blood cytometry measurements and peripheral blood gene expression measures. RNA-Seq data for 241 participants from the BIODEP

study was generated from PBMCs using TruSeq Stranded mRNA kit libraries resulting in 75bps paired-end reads, and sequenced on Illumina HiSeq 4000. Average sequencing depth was 54.5 million reads. For additional information on PBMC isolation and RNA sequencing methodology see Cole et al (1).

2) **Le dataset:** Control and MDD participants (meeting DSM-IV-TR criteria) for the study (2) were recruited from Oklahoma, USA. MDD severity was ascertained using Montgomery-Asberg Depression Rating Scale (MADRS). RNA-Seq data for 160 participants were generated from PBMCs, libraries for sequencing were prepared using TruSeq Stranded mRNA kit and the resulting 150 bps paired-end reads were sequenced on Illumina HiSeq 3000. The average sequencing depth was 30 million reads. Access to raw data was established in collaboration with Dr Brett A. McKinney, University of Tulsa, USA.

3) **Mostafavi dataset:** Participants for this study (3) were recruited from the USA. MDD patient screening was performed using the Composite International Diagnostic Interview (CIDI), evaluating scores from DSM-IV, Patient Health Questionnaire (PHQ-9), 7-item scale Generalized Anxiety Disorder (GAD-7), and a family history screen for MDD, bipolar disorder and suicide. Whole blood samples from 922 individuals were used to generate RNA-Seq data. Libraries were prepared using Illumina TruSeq kit with poly(A) selection and sequenced on Illumina HiSeq 2000. 50-51bps single-ended reads were generated with an average sequencing depth of 70 million reads. This dataset was obtained through application to the National Institute of Mental Health (NIMH) Genetics Repository (https://www.nimhgenetics.org/), study 88.

4) **dbGaP:** Microarray data for control and MDD samples were obtained through application via the dbGaP web portal (https://www.ncbi.nlm.nih.gov/gap/ddb/), study accession: phs000486.v1.p1. The study consists of two cohorts: the Netherlands Study of Depression and Anxiety (NESDA), and the Netherlands Twin registry (NTR). CIDI screening followed by DSM-IV evaluation was used for MDD case ascertainment. Total RNA was isolated from whole blood samples using the PAXgene Blood RNA MDx kit protocol with the BioRobot Universal System (Qiagen), and samples were hybridized to Affymetrix U219 array plates. For additional information on sample extraction and microarray processing see Wright et al (4). NCBI dbGaP gave approval for this re-analysis under the project number #28948.

5) **HiTDiP:** CNT and MDD participants were recruited as part of the GlaxoSmithKline-High-Throughput

Disease-specific target Identification Program (HiTDiP). RNA was isolated using the standard PAXgene protocol on BioRobot 8000 (Qiagen), and samples were hybridized to Affymetrix U133 plus 2.0 array plates. Raw microarray data for this study was obtained from NCBI GEO with accession number GSE98793 (https://www.ncbi.nlm.nih.gov/geo/query/acc.cgi?acc=gse98793).

**Sample processing**

Five MDD case-control datasets: three RNA-Seq (BIODEP, Le, and Mostafavi) and two microarray (dbGaP and HiTDiP), met criteria and were analyzed. Raw RNA-Seq .FASTQ files and microarray .CEL files were processed to generate gene and/or transcript expression values. Raw data for samples were analyzed following the same processing pipeline as BIODEP for RNA-Seq datasets and the same pipeline as dbGaP for microarrays datasets, as only the expression matrix was available for dbGaP, and BIODEP was previously processed in-house.

Covariates were variably available across datasets. To maintain homogeneous processing across datasets, only covariates that were available across all five datasets - batch (i.e. study center, sequencing plate ID), sex, and age - were used for transcriptomic models. MDD case status was significantly associated with sex, batch, age, BMI, and time of blood draw in dbGaP; and with sex, BMI, and smoking in Mostafavi (**Table S1**).

**RNA-Seq data processing**

To minimize the influence of processing pipeline differences on gene and transcript expression quantification, raw data for each dataset was processed using a common pipeline, and default values were used unless specified (**Figure S2**). A processed gene count matrix was available for our in-house dataset BIODEP (75bps paired-end reads), and the same processing steps were applied to the other two RNASeq datasets: Le (150bps paired-end reads) and Mostafavi (50bps single-end reads). Read quality was assessed using FastQC (version (v) 0.11.9) and visualized using MultiQC (v1.11.dev0). The Le raw reads showed presence of adaptor sequences, and therefore, read trimming was performed using cutadapt (v1.9) ensuring that the trimmed sequence has a minimum quality score of 30 and final length of at least 50bps (same parameters as that used in BIODEP processing). For all the three datasets, reads were aligned to the GRCh38 (v84) genome (GRCh38 version 84 primary assembly FASTA file and GTF file: http://www.ensembl.org/info/data/ftp/index.html/) using STAR (v2.5.2b), and uniquely mapped reads were quantified per gene with featureCounts (v1.5.1) with GRCh38 v84 GTF file as reference. Gene IDs were annotated with gene names from Ensembl (GRCh38 v84) using biomaRt (v2.46.3) in R. Only genes from autosomal chromosomes 1-22, sex chromosomes X and Y and mitochondrial chromosome (MT) were retained for further analysis.

For transcript-level analyses, transcriptome alignment was performed using salmon (v1.4.0) with transcripts from both protein-coding (cDNA FASTA) and noncoding (ncDNA FASTA) regions as reference (GRCh38 v84: http://www.ensembl.org/info/data/ftp/index.html/) to allow for detection of noncoding transcripts in the samples, a small subset of which have poly(A) tails. The genomic sequence information was used as decoy during transcript alignment to reduce erroneous mapping of reads, for example, from unannotated genomic loci with a similar RNA sequence to annotated transcripts. We opted for parameters that corrected for sequencing biases - arising from preferential sequencing of read fragments starting with certain nucleotide motifs; GC biases in the data to improve transcript quantification; and resampling was performed 100 times to capture technical variance in transcript count estimation.

**Microarray data processing**

For microarray datasets, processed gene counts (but not raw data) were available for dbGaP and, therefore, the same processing steps applied by the dbGaP authors were used for the HiTDiP analysis to standardize processing. Raw .CEL files were read and normalized using affy (v1.68.0). The normalization performed included: background correction of intensity values using Robust Multi-array Average (RMA) method, quantile normalization to minimize any technical differences between experiments, and $\log_2$ transformation of expression values. Probes were annotated with the hgu133plus2 database using AnnotationDbi (v1.52.0); unannotated probes and probes mapping to multiple genes were removed.

**Sample filtering: RNA-seq**

Samples without any metadata or metadata specifying that the patient requested withdrawal from the study were removed. Additionally, if RNA integrity number (RIN) information was available, samples were filtered to have RIN $\geq 8$. Diagnostic principal component analysis (PCA) and multidimensional scaling (MDS) plots were used to identify outlier samples. To evaluate concordance between recorded sex labels and expected sex-specific gene expression, average expression of chromosome Y genes, and hierarchical clustering using average *XIST* gene expression (located on chromosome X) were performed. We expect genes corresponding to chromosome Y to be expressed only in people assigned male at birth, and *XIST* gene expression to be high only in people assigned female at birth as the *XIST* gene in male chromosome X is normally inactive. Samples for which recorded sex did not match genome-based biological sex categorizations were removed from further analysis.

**Sample filtering: microarray**

Samples without corresponding metadata were removed. Furthermore, for a subset of dbGaP samples, samples from the same individual were collected twice but at different time points. Only one of the time points had corresponding cell count information, and so was retained for further processing. Normalized gene expression values were evaluated using arrayQualityMetrics (v3.46.0) in R, and outliers were identified using three methods: if the sum of the Euclidean distances of an array to all other arrays was exceptionally large; if the signal intensity distribution of an array was comparatively different from the intensity distribution of the pooled arrays as determined by the Kolmogorov-Smirnov statistic; and MA plots were prepared between an array of interest and a 'pseudo' array computed using the median of all arrays evaluated, and the joint distribution of M (log intensity difference) and A (mean average intensity) was evaluated using Hoeffding's statistic $D$ (threshold of $D < 0.15$). Additionally, as for the RNA-Seq data, any chromosome Y gene and XIST expression-based inconsistencies with recorded sex were identified and removed. Final sample sizes after outlier removal are shown in **Table 1.**

**Covariate selection**

For categorical covariates such as study center, sex, and condition, MDS plots were used to visualize covariate contribution to sample cluster separation. Covariates were also assessed for association with MDD using a t test for continuous variables and a Chi-squared test for categorical variables.

**Adjusting for sample cellular composition using assayed counts and cellular deconvolution**

To account for gene expression variation due to cell composition differences in bulk RNA-Seq data, and between PBMC and whole blood samples, cell counts were included in gene and transcript expression models. Cell count information was available for a majority of BIODEP participants (B cells, CD4+ and CD8+ T cells, monocytes, natural killer (NK) cells) and for a subset of dbGaP samples (lymphocytes, neutrophils, eosinophils, basophils, monocytes). For all datasets except BIODEP (where we used cytometry counts), cell count proportions used in expression models were estimated from gene expression data using xCell (v1.1.0) to estimate per sample enrichments of B cells, CD4+ T cells, CD8+ T cells, NK cells, monocytes and (additionally for whole blood samples) eosinophils, basophils and neutrophils. xCell uses cell type-specific gene signatures curated from multiple sources to determine the level of cell type-specific enrichment in a gene expression dataset.

To run xCell on RNA-Seq data, raw gene counts were converted to Reads Per Kilobase of transcript per Million (RPKM) values, which account for gene length differences, using the RPKM function from edgeR (v3.32.1), with gene length information derived from the ensembldb (v2.41.1) package. For gene IDs mapping to the same gene name, the longest gene length was used for the calculation. Transcript Per Million (TPM) values - expression values which are comparable across samples, were calculated from RPKMs as follows:

$$t\left(\frac{t(RPKM)}{colSums(RPKM)}\right) * 1e^{6} \qquad \text{[Eq 1]}$$

For microarray data, RMA normalized counts were used with xCell as per the authors recommendation.

For datasets derived from isolated PBMCs, cell enrichment values were calculated for B cells, CD4+ T cells, CD8+ T cells, monocytes, and NK cells. For datasets derived from whole blood samples, we also calculated enrichment of additional cell types present in whole blood not PBMCs (basophils, eosinophils and neutrophils). We validated the xCell results by comparing (using Spearman correlations) xCell generated values to original cell count values for dataset subsets with cell count information. Basophils were found to have poor correlation between original cell counts and xCell generated values, and therefore, were not included in the final cell correction model (**Figure S4**). We note that xCell is not strictly a deconvolution tool but calculates cell type enrichment within samples and is appropriate for comparing cell type enrichments between samples. We focused on xCell as it outperforms other tools in detection of minor cell types such as eosinophils which were important for our sample types (5).

**Differential gene and differential transcript models**

See **Figure S2** and **Supplementary Methods** for details of differential gene expression (DGE), differential transcript expression (DTE) and differential transcript usage (DTU) pipelines. For DGE and DTE, we considered only features (genes or transcripts) present in all included datasets, to focus on features for which the meta-analysis was best powered (11,845 genes for DGE; 38,877 transcripts from 14,186 genes for DTE). Duplicate gene names were filtered to retain the entry with the higher $\log_2$ expression value ($\log_2$CPM). Features for the non-sex-stratified analyses were modelled as follows, where cell counts used were those assayed using flow cytometry (for BIODEP) or estimated using xCell (all other datasets); for this main model, BIODEP participants without cytometry data were excluded. For PBMC samples, features were modelled as follows ($\beta$ indicates regression coefficients):

$$\text{counts} \sim \beta_0 + \beta_1*\text{condition} + \beta_2*\text{age} + \beta_3*\text{sex} + \beta_4*\text{batch} + \beta_5*\text{B cells} + \beta_6*\text{CD4+ T cells} + \beta_7*\text{CD8+ T cells} + \beta_8*\text{monocytes} + \beta_9*\text{NK cells} \quad \text{[Eq 2]}$$

Models for whole blood samples additionally included neutrophil and eosinophil counts but not basophils due to the poor correlation of xCell-estimated basophils with recorded basophils (**Figure S2**):

$$\text{counts} \sim \beta_0 + \beta_1\text{*condition} + \beta_2\text{*age} + \beta_3\text{*sex} + \beta_4\text{*study centre} + \beta_5\text{*B cells} + \beta_6\text{*CD4+ T cells} + \beta_7\text{*CD8+ T cells} + \beta_8\text{*monocytes} + \beta_9\text{*NK cells} + \beta_{10}\text{*eosinophils} + \beta_{11}\text{*neutrophils} \quad \text{[Eq 3]}$$

Additional sensitivity DGE analyses included (a) the above models without any cell counts included; (b) the above models with BMI as an additional covariate, for the three datasets where BMI was available (BIODEP, Mostafavi, and dbGaP) datasets; (c) the above models with an additional term for (xCell-)estimated platelet counts for the whole blood (Mostafavi, dbGaP, and HiTDiP) datasets.

DTU analysis of MDD case-control studies was conducted using DEXSeq (v1.36.0) to conduct the differential transcript analysis (performed at the transcript-level), see later in this document. DEXSeq fits a generalized linear model (null model + the interaction term condition:transcript) for each gene and compares it to the null model. For PBMC samples this was:

count ~ sample + exon + study centre:exon + age:exon + sex:exon + B cells:exon + CD4+ T cells:exon + CD8+ T cells:exon + monocytes:exon + NK cells:exon [Eq 4]

Null model for whole blood samples:

count ~ sample + exon + study centre:exon + age:exon + sex:exon + B cells:exon + CD4+ T cells:exon + CD8+ T cells:exon + monocytes:exon + NK cells:exon + eosinophils:exon + neutrophils:exon [Eq 5]

Following DTU analysis, stageR (v1.12.0) was used to generate adjusted transcript *P*-values while controlling for overall gene-level FDR.

Sex-stratified analyses were conducted without the ‘sex’ variable in the models shown in Equations 2-5.

**Differential gene expression (DGE) analyses (per-dataset)**

To allow for standardized processing, covariates common to all five MDD case control datasets were used for differential expression analyses: age, sex, batch (sequencing center, plate, etc.), and cell count enrichment values (i.e. those subsets listed above excluding basophils, which did not show good correlation between estimated and observed cell counts in the datasets for which both were available, see **Figure S4**).

**RNA-Seq data:** Read counts were processed using functions from the R package edgeR (v3.32.1): genes with low expression were filtered out (< 10 read counts in the smallest group between control or MDD), and library size normalization was performed. These RNA-seq counts were transformed for linear modelling using the limma function limma-voom, and the limma-voomwithQualityWeights function (6) was used to model heteroscedasticity and down-weight observations from more variable samples to improve power.

**Microarray data:** RMA normalized gene counts were assessed for differential gene expression between MDD cases and controls. A linear model was fit for each gene using the limma function lmfit, and then eBayes was used to calculate a moderated t-statistic, which used information from all input genes to calculate variance.

In both cases, gene expression models were as per Eq.2 and Eq.3 above and conducted using the R package limma (v3.46.0). Only genes common to all 5 datasets were included in analyses. Results were FDR corrected using the BH method and DGE was called at a threshold of FDR<0.05.

**Differential transcript expression (DTE) analysis (individual dataset results)**

DTE analyses were performed for the three MDD case-control datasets which used RNAseq, as transcript-level analyses are not possible from microarray data. Salmon-generated transcript counts were read and annotated via the tximeta package (v1.8.5) (7). DTE analysis was performed for the three RNA-Seq datasets using swish from the fishpond package (v1.6.0) (8). Swish calculates the test statistic using a Mann-Whitney U test with a pseudo-random generator (drawn from a uniform distribution of (0, 0.1]) to break ties. Transcript counts were corrected for transcript length and sequencing depth, and transcripts with low expression were removed (the same criterion used in the DGE processing). The same covariates used in DGE analysis were also used for DTE, however, the covariates were corrected using removeBatchEffect from limma per the author's recommendations. DTE results were identified at an FDR threshold of 0.05.

**Differential transcript usage (DTU) analysis (individual dataset results)**

Our pipeline for DTU analysis of MDD case-control studies was based on three packages: DEXSeq (v1.36.0) to conduct the differential transcript analysis (performed at the exon-level), stageR (v1.12.0) to perform gene-wise correction of transcript *P*-values, and IsoformSwitchAnalyzeR (ISAR) (v1.12.0) to annotate and visualise the isoform structure and any protein domains within its sequence to enable inference of isoform function. ISAR incorporates DEXSeq for DTU analysis and offers superior functions for isoform switch visualization. We wrote bespoke wrappers and modified ISAR code to allow for parallelization and customized outputs. We imported salmon-quantified transcript counts, added transcript annotation and coordinate information using GRCh38 v84 GTF and FASTA files as references (same as the references used during transcriptome alignment using salmon); filtered transcripts with low expression (minimum average gene expression (FPKM) in both conditions >3 and minimum transcript count >0 in both conditions); and removed genes with only one transcript. The output was stored in ISAR's 'switchlist' format – a compatible format for storing annotations and other gene and transcript information for downstream analysis. DEXSeq was used to calculate dispersions (using a negative binomial distribution) and test for differential exon usage with parallelization enabled and to calculate per gene *Q*-values (FDR adjusted *P*-values). stageR was used adjust transcript-level p-values for overall FDR (OFDR).

DTU results were not meta-analyzed as the modelling approach entails a highly non-uniform p-value distribution, precluding weighted Z-score meta-analysis. For the three RNA-Seq datasets for which DTU could be analyzed, 0 genes were DTU in the BIODEP dataset; 118 genes were DTU in the Le dataset and 0 genes (not cell corrected as cell corrected results did not converge) were DTU in the Mostafavi dataset (see Zenodo https://doi.org/10.5281/zenodo.15290507). *GPX1*, which showed DGE in the female-only meta-analysis, also showed differential transcript usage in the Le dataset in the sex-pooled (gene-level FDR= 1.2e-7) and female-only (gene-level FDR= 0.002) analysis. The other 40 DGE genes (FDR<0.05) from the main meta-analyses did not show evidence of DTU.

**Weighted Z-score meta-analysis**

A weighted (weighted by sample size) Z-score method was used to combine the *P*-values using metapro: https://github.com/unistbig/metapro (9). Direction of effect was evaluated using the sign of $\log_2$FCs (a measure of effect size here). Input weights reflected sample size for each dataset. Resulting p-values were corrected for multiple testing using the BH method and significant results were called at FDR<0.05.

Per-dataset gene and transcript differential expression results were meta-analyzed using a bias and inflation-corrected weighted Z score method. Z scores were calculated from raw DGE or DTE P-values and the sign of the $\log_2$FC values was used to determine the sign of the Z scores for each gene. Genes analyzed for DGE were those present in all five datasets; transcripts analyzed for DTE were those present in all three datasets with transcript-level data. Standard methods to control for p-value inflation used in GWAS are not suitable for transcriptomic studies; we used R package BACON to calculate bias and inflation values representing deviation of the mean and standard deviation, respectively, from the theoretical null distribution (10). BACON is a statistical method to correct for bias and inflation in p-values. It uses a Bayesian method - a Gibbs sampling algorithm, to fit a three-component normal mixture to a set of test statistics (for example, DGE P-values). One of the resulting components is an empirical null distribution whose parameters represent bias and inflation estimates for the dataset in question. The other two components capture the fraction of true associations in the data which helps address overestimation of the inflation parameter known to occur in data with moderate amounts of true associations. For each dataset (*i*), bias and inflation values calculated using these Z scores were used to generate inflation- and bias-corrected Z scores (Eq 6) and the per-dataset modified Z scores ($x_i$) were used to calculate the meta-analytic weighted Z scores (Eq 7):

$$x_i = (Z\ score - bias)/inflation \qquad \text{[Eq 6]}$$

$$Z_{weighted} = \sum^{n}(x_i * w_i)/\sqrt{\text{sum}({w_i}^2)} \qquad \text{[Eq 7]}$$

Where $w_i$ is the effective sample size for dataset *i* calculated as:

$$w_i = 4/\sqrt{[(1/\text{number of cases}) + (1/\text{number of controls})]} \qquad \text{[Eq 8]}$$

Meta-analytic *P*-values were calculated from weighted Z-scores and were corrected for multiple testing using the BH method. Significant results were called at an FDR threshold of 0.05. P-values generated from the DTU pipeline are not expected to be uniform and therefore cannot be meta-analyzed using the weighted Z score method.

Bacon inflation estimates were lower where the model included cell counts, compared to the model without cell counts (main $\lambda$=1.21 vs uncorrected $\lambda$=1.31), and slightly lower for the sensitivity analysis that additionally included BMI, compared to the main analysis (main $\lambda$=1.21 vs $\lambda$=1.14 with BMI), suggesting that accounting for cellular composition and BMI helps to reduce heterogeneity; this remained true when we reperformed the meta-analyses with matched sample sizes for all models. When we reperformed the meta-analyses without correcting for cellular composition in each dataset model, the only DE genes were *CD247* (decreased in MDD in sex-pooled analysis) and *IL2RB* (deceased in MDD in male-only meta-analysis), with no DGE in the female-only meta-analysis.

**Comparison with depression proteome**

To compare our results to MDD-associated proteins, we obtained results from a recent analysis of the MDD-associated plasma proteome where 1,463 proteins were assayed using Olink technology and tested for association with electronic health record-defined depression in 54,219 participants from UK Biobank. Results were obtained from Supplementary Table 10 of Daskalakis *et al.* (11)

**Sensitivity meta-analyses**

To test the sensitivity of our results to adjustment for BMI, for those cohorts where BMI was available (BIODEP, Mostafavi, dbGaP), we also performed a meta-analysis additionally corrected for BMI (**Table S8**). For the 40 DGEs significant in our main meta-analyses, 20 (including *ACRBP* and *CDIPT* and the genetically prioritized gene *GPX4*) remained genome-wide significant in the BMI-adjusted model. For 31 of the 40 DE genes, the meta-analytic direction of effect was the same following BMI adjustment. Our core biological findings also remained significant using the BMI-adjusted meta-analysis results: the same cell cycle and mitochondrial pathways were significantly downregulated in the BMI-corrected meta-analysis (**Figure 2A**). Cell origin analysis again prioritized CD8+ T and NK cells (CD8+ TEM coef=0.005; FDR<0.001; NK2 coef=0.006, FDR<0.001), and additionally implicated memory regulatory T cells as contributors to the DGE profile (coef=0.006; FDR<0.0001, **Figure S7**).

For the leave-one-out meta-analyses, excluding each dataset in turn prior to meta-analysis, for the 2 differentially expressed genes in our main sex-pooled analysis, both genes showed the same direction of effect and were significant at unadjusted P<0.05 in four out of five LOO analyses (all except LOO dbGaP).

**Enrichment analyses**

Pathway or gene set enrichment analyses (GSEA) were performed using gene lists from the Reactome database v75, a set of curated human biological pathways (12) and the ImmuneSigDB database (13), accessed via MSigDB, which contains 4,872 gene sets identified from 389 immunological studies, capturing the transcriptomic effects of chemical and genetic perturbations of the immune system (14, 15). GSEA was performed using meta-analyzed DGE (for ImmuneSigDB and Reactome) or DTE results (for Reactome only; DTE is less interpretable with respect to MSigDB which is generated empirically from gene-level experimental results). Duplicate gene names were filtered to retain the entry with the higher $log_2$FC. For DTE, transcripts were grouped by their corresponding genes and the transcript with the smallest P-value was chosen to represent the gene. GSEA was performed on genes ranked by signed DGE/DTE P-values, where:

$$signed\ P_{value} = [-log_{10}(P_{value}) * sign(log_2(Fold\ Change))] \quad \text{[Eq 9]}$$

Gene set enrichment analysis (GSEA) was performed using the GSEA function from clusterProfiler (v3.18.1), where 10,000 permutations were used to compute P-values, and significance was evaluated using an FDR (BH method) threshold of 0.05. We also performed a GSEA analysis focused on T cell activation states by reprocessing an human *ex vivo* dataset which assayed the transcriptomic response to stimulation of naive and memory CD4 T cells with varying concentrations of anti-CD28 and anti-T cell receptor (16). We generated gene sets from these publicly available differential expression profiles using a method matching ImmuneSigDB as follows: the subset of genes with FDR < 0.02 were ranked per their signed P-values (Eq 8). For each T cell activation state, 'up' and 'down' gene sets were created using the top 200 upregulated and downregulated ranked genes, respectively (or fewer genes if fewer than 200 were FDR < 0.02).

**DTE pathway enrichment results**

DTE pathway enrichment analyses (see **Figure S5**) highlighted decreases in pathways related to cell cycling (G1/S-specific transcription, NES=-2.01, FDR=0.02), but additionally detected increases in multiple pathways suggesting increased translation initiation (formation of a pool of free 40S ribosomal subunits, NES=2.078, FDR=0.003 and GTP hydrolysis and joining of the 60S ribosomal subunit, NES=2.077, FDR=0.003). There was also an increase in GCN2-response to amino acid deficiency, a pathway closely linked to control of translation, with translation-initiation pathways consistently increased in both males and females**.** In the gene-level analysis these pathways were not detected as differentially expressed even at unadjusted $P<0.05$.

**Transcriptome-wide association (TWAS) integration**

To investigate the relationship of our observed results from patient cohorts with genetically predicted DGE, we tested the concordance of our results with those of a recent transcriptome-wide association study (TWAS). This TWAS estimated the blood gene expression changes predicted to be associated with depression by integrating genetic risk variants for the disorder with quantitative trait loci (QTL) data on the genetic predictors of gene expression in blood. Only two of the 1,550 genes that were genetically predicted to be differentially expressed in blood by TWAS at adjusted P < 0.05 showed concordant differential expression in our real-world case-control meta-analysis: *GPX4* and *GYPE* were decreased in blood in females with MDD ($Z_{meta}$=-3.79; FDR<0.05 and $Z_{meta}$=-3.97, FDR<0.05 respectively), as genetically predicted, with both genes showing a trend to decrease (unadjusted P<0.05) in the sex-pooled meta-analysis. Notably, the whole blood eQTLs used in the TWAS we drew on were not corrected for cellular composition; to test whether the observed discordance between genetically-predicted and observed DGE was simply due to our inclusion of cell counts in DGE models, we generated mega-analytic results uncorrected for cell composition and re-examined DGE-TWAS concordance; again, there was little concordance between observed and genetically predicted DGE (**Figure S4D**).

**Rank-rank hypergeometric overlap analysis**

To compare DGE results (including non-genome wide significant genes) between male and female samples, and to compare DGE results to TWAS-predicted DGE, we conducted a threshold-free rank-rank hypergeometric overlap (RRHO) analysis using the R package RRHO2 (v1.0) (17). Briefly, two gene lists of interest are ranked by their degree of differential expression represented by their P-value and the sign of their effect size (here, $-\log_{10}$(P-value) * sign($\log_2$FC) for DGE and $-\log_{10}$ (TWAS P-value) * sign (TWAS Z score) for TWAS-predicted DGE) and are plotted along the x and y axes from most upregulated to most downregulated. For each combination of cut-offs along the x and y axes, subsets of gene lists from the outer corner of the quadrant to cut-off point are calculated, and the significant number of overlapping genes is determined using the hypergeometric distribution. This analysis was conducted using the RRHO2_initialise function using default parameters except log10.ind = TRUE. Significant overlap for each (x,y) combination, captured as a 2x2 enrichment table, is used to calculate the hypergeometric p-value. The resulting hypergeometric p-values ($-\log_{10}$ scale) were visualized using the RRHO2_heatmap function with default parameters. The resulting RRHO heatmaps show hypergeometric p-values ($-\log_{10}$ scale) visualizing the extent of concordance (top right and bottom left quadrants) and discordance (top left and bottom right quadrants) between two disorder-association gene lists. We used this RRHO method to visualize the concordance and discordance of genetically-predicted vs. observed MDD-associated DGE, and of male vs. female observed MDD-associated DGE.

**Curation of MDD-associated focused gene lists involved in core biological processes**

Alongside these data-driven analyses, we also performed more focused analyses motivated by the existing literature on pathways likely to be implicated in MDD. The gene groups evaluated, and their sources are listed below:

1) **Mitochondrial chromosome:** Chromosome information for genes was extracted from Ensembl using biomaRt. Genes were filtered to entries from the mitochondrial “MT” chromosome.
2) **Nuclear mitochondrial:** A list of genes located in the nuclear chromosome with protein products known or predicted to localize in the mitochondria were curated from the Integrated Mitochondrial Protein Index (IMPI) via the MitoMiner database v4 (18).

3) **Ribosomes:** Gene descriptions corresponding to gene names were extracted via biomaRt, filtered to include "ribosome" genes, and filtered to remove any genes from the chromosome MT.
4) **Replication-dependent and replication-dependent histones:** Gene descriptions corresponding to gene names were extracted via biomaRt and filtered to include "histone" genes. Information on replication types for histones (replication-dependent, replication independent or pseudogenes) was curated from NCBI gene summaries (https://www.ncbi.nlm.nih.gov/gene).
5) **Cell cycle:** Genes involved in a specific phase of the cell cycle were downloaded from Cyclebase v3 (19).
6) **Oxidative phosphorylation (OXPHOS) electron transport chain (ETC):** Genes involved in ETC corresponding to the Reactome pathway "The citric acid TCA cycle and respiratory electron transport".

For pooled, male and female samples (3 separate analyses), we tested the significance of the median log fold change of the gene set in MDD vs. controls using a two-sided permutation test to test for robustness of observed gene set differences against chance expectations. This tested, for example, whether replication-dependent histones were on average increased or decreased in MDD compared to controls. For each gene set, we tested whether its median expression differed from zero, with permutations (n=1000) run within each dataset and combined via meta-analysis (weighted Z-score method). This approach differs from standard GSEA as it captures consistent gene group-level expression changes and adjusts for baseline gene expression, enhancing robustness but requiring intensive computation.

For permutations, the replacement set was chosen from the list of genes that did not belong to any of the 13 gene group under investigation. A problem with random selection from this list is that genes with overall lower expression are less likely to be detected as differentially expressed. To account for this bias, we ensured that the substitute genes (selection performed without replacement) had similar expression values to the gene list being tested. Replacement genes were selected to match baseline expression of the genes in the gene list, choosing genes randomly from a bin of replacement genes with a maximum difference in baseline expression of 0.1 $\log_2$CPMs. For genes with minimum replacement options this bin was expanded to ±4 $\log_2$CPMs. For each of the matched replacement sets (N=1000), median $\log_2$FC was calculated for pooled samples, females and males to generate a distribution of median $\log_2$FC values. The observed median $\log_2$FC of the gene set ($median_{actual}$) was evaluated against this distribution and p-value for the permutation test was calculated as:

$$P = \frac{total\left(abs(median_{perm}) \geq abs(median_{actual})\right)+1}{1000+1} \quad \text{[Eq 10]}$$

This was performed separately for each dataset. The per-dataset p-values were meta-analyzed using a weighted z-score method, and adjusted for multiple testing (N=13 gene sets) using the BH method.

**Consensus weighted gene co-expression network analysis (WGCNA)**

We sought to identify modules of co-expressed genes consistently dysregulated in MDD, while accounting for sample cellular composition. To do this, we used consensus WGCNA, which generates a gene co-expression network consistently present across multiple datasets.

Gene-gene correlations (similarities) used construct the gene co-expression network were calculated using Biweight Midcorrelation (bicor) - a median-based approach that is more robust to variability in gene expression compared to mean-based Pearson correlations. The soft-thresholding power parameter (*ß*) which produced networks with scale-free topology fit of $R^2 > 0.8$ and mean connectivity between 30-100 in all five datasets were chosen for consensus WGCNA. If soft-thresholding power did not reach these criteria for an individual dataset (reflecting noise), the soft-thresholding power at the point of inflection of $R^2$ values was chosen instead. This method identified optimal soft-thresholding powers of 12 for female and 10 for male consensus networks.

Similarities calculated between gene pairs were used to construct a similarity matrix with rows and columns representing individual genes. Gene networks quantified by positive gene-gene correlations have been shown to be more robust than networks quantified using positive and negative gene-gene correlations. Therefore, a signed WGCNA network was constructed, which considers negatively correlated genes or nodes as unconnected. In signed networks, highly correlated genes are given more importance than genes with lower correlations by scaling the similarity matrix by a soft-thresholding power (*ß* ≥ 1) to calculate adjacencies:

$$aij = \left|0.5 + 0.5\left(bicor(xi, xj)\right)\right|^{ß} \quad \text{[Eq 11]}$$

Here, $x_i$ and $x_j$ represent gene pairs being compared, where high negative correlations of -1 would have 0 adjacencies and would not contribute to edge calculations.

A Topological Overlap Matrix (TOM) was used to construct a weighted network of highly proximal or interconnected genes, where edges for each node or gene pair represents the sum of adjacencies of the gene pair with other genes in the network:

$$TOM_{ij} = \frac{\left|a_{ij} + \Sigma_{u \neq i,j}\left(a_{iu} \times sign\left(cor(x_i, x_j)\right)\right) \times \left(a_{uj} \times sign\left(cor(x_i, x_j)\right)\right)\right|}{\left(k_i, k_j\right) + 1 - \left|a_{ij}\right|} \quad \text{[Eq12]}$$

Here, $TOM_{ij}$ – proximity between genes *i* and *j* – represents an element of the TOM matrix, *a* represents adjacencies, *x* represents gene expressions, and $k_i$ represents connectivity of node *i* which is the sum of adjacencies between a node and every other node.

A dissimilarity matrix (1-TOM) was used as input to hierarchical clustering by the average-linkage method to construct gene network dendrograms. In the average-linkage method, Euclidean distances between clusters are computed as average distances between each pairwise cluster component. The dendrograms were cut into closely related gene groups (defined by the dendrogram height: representing the distance metric - the higher the point of first connection between two clusters, more the dissimilarity between the clusters) or modules with user-specified parameters: deep split (*d*) which controls how sensitive the algorithm for module splitting is to relatedness between modules, minimum module size (*m*) to control the minimum number of genes in a cluster for module identification, and cut height (*h*) specifying the dendrogram height at which similar modules should be merged with each other. These algorithms were accessed via the R package wgcna (v1.70-3).

Log expression values for all 5 datasets were corrected for study center, age, and cell counts using limma removeBatchEffect and filtered to retain only genes common to all datasets ($n$ = 11,857). Separate male and female consensus WGCNA networks were constructed as follows: consensus gene-gene similarities were identified by first calculating individual topological overlap matrices (TOMs) for each dataset (Eq. 4) and then combining the TOMs such that for each element of the TOM, the minimum TOM across datasets was chosen to generate the final consensus TOM matrix. This is a conservative method in that it retains only gene-gene connections present in all datasets. Consensus WGCNA was performed using the blockwiseConsensusModules function in wgcna with maxBlockSize set to 20,000 to allow all genes to be processed together and parameters $d$ = 2, $m$ = 75 and $h$ = 0.25 specified.

Although the consensus modules represent consistent gene groupings across datasets, the strength and pattern of co-expression may still vary between datasets. To capture this, we used module eigengenes to represent overall module expression. Consensus module eigengenes are the per-module first principal components of the gene expression matrix for genes in that consensus module. Gene reassignments and module trimmings were also performed by removing genes that had a correlation of less than 0.3 (default value) with their respective module eigengenes, and further, modules without at least $m$/3 gene-module eigengene correlations of greater than 0.5 were disbanded and the genes were considered unassigned (by default, grey colored modules).

To identify hub genes in a module in each dataset, kME values were calculated as the correlation (bicor) between a gene's expression and the consensus module eigengene. Higher kME values indicate highly connected genes within a module. Average kMEs across the 5 datasets, weighted by sample numbers of the individual datasets, were ranked to identify hubs genes for each consensus module i.e. genes with expression profiles that are highly representative of the expression of the module. Associations between consensus module eigengenes and clinical phenotypes were tested using the wgcna

Spearmancorrelation function. Significant associations were identified using a student asymptomatic *P*-value < 0.05.

Per-module biological process gene ontology (GO) term over-representation analyses were performed using a one-sided Fisher exact test via the enrichGO function in clusterProfiler (v3.18.1) with a BH adjusted *P*-value threshold of 0.05. For each module, the significant GO term (FDR $P < 0.05$) with the highest gene ratio was selected as the module representative. Gene ratios were calculated as the number of module genes associated with a GO term divided by the number of genes in all the modules associated with the same GO term. Eigengene networks (showing Pearson correlations between different module eigengenes) were constructed to visualise inter-dataset similarities in the consensus WGCNA modules; module preservation scores were used to capture inter-dataset differences in these eigengene networks.

**Enrichment analysis for WGCNA consensus modules**

Significant enrichments for 249 biological process GO terms in females and 325 terms in males were found for consensus modules and the most enriched module (by GeneRatio: number of module genes in the gene set/number of genes in the gene set for genes with FDR < 0.05 or $P < 0.05$ if no significant terms were identified) were selected as the module's characteristic pathway. We note that the hub gene for a given module is not necessarily a part of the most enriched pathway.

**Module preservation scores**

Although consensus modules were identified across the five MDD case-control datasets, the eigengenes corresponding to these modules were data-specific as intra-dataset gene expression profiles were different. To visualise inter-dataset similarity of the consensus WGCNA modules, eigengene networks (showing Pearson correlations between different module eigengenes) were constructed. In eigengene networks, nodes are module eigengenes and edges represent correlations between them.

To assess how well these networks matched across datasets, we computed mean preservation scores (*Pres*), which quantify how similar each module's connectivity pattern is across datasets (20). *Pres* scores quantify inter-dataset differences in eigengene networks by calculating how well the modules in a reference set are preserved in the test set using genes common to the two sets:

$$Pres\_ij=1-[\max(A_{ij}(1),A_{ij}(2)) - \min(A_{ij}(1),A_{ij}(2))] \quad \text{[Eq 13]}$$

Here, $A_{ij}$ refers to adjacencies calculated using correlations between eigengenes *i* and *j* for dataset (1) and (2). Mean preservation (*D*) was identified by (1) calculating the mean of preservation values for an eigengene to all other eigengenes, and then (2) calculating the mean of all per-module mean preservation values from step 1.

Overall, consensus modules were highly preserved, with high mean preservation scores in both male and female networks (**Figure S10**), and no clear difference in preservation between RNA-seq and microarray datasets, indicating robust cross-platform reproducibility.

**Differential expression of WGCNA modules**

Five batch-corrected MDD case-control data (corrected for study center, age, and cell counts) were processed to retain only common genes (*n* = 11,857). Dataset processing affects WGCNA results, and therefore, datasets were processed as similarly as possible. As the VST method of data normalization is applicable only to RNA-Seq data, and here we also analyze microarray data, VST was not used and instead $\log_2$ transformed microarray intensities, and $\log_2$ transformed RNA-Seq counts corrected for library normalization were used for WGCNA. Both control and MDD samples were used to construct networks to increase sample size, and to avoid loss of information on MDD-specific module. The association between MDD and module eigengene scores was tested by student's t-test; p-values from these t-tests were meta-analyzed across datasets using a weighted Z-score method.

A consensus module with *PUF60* as the top hub gene was detected in both the male consensus network (module ME3, 445 genes) and female consensus network (module ME5, 279 genes), with 180 genes in common between the male and female *PUF60* module. This module was upregulated in MDD in both the female and male WGCNA meta-analyses (see **Figure 3**). For the female WGCNA analysis, the hub genes of the other modules upregulated in MDD (ME7, ME4 and ME2) were, respectively, *GMPR,* an enzyme that reduces guanosine monophosphate to inosine monophosphate; *COX4I1*, an enzyme in the mitochondrial respiratory chain, and *UBXN1,* a negative regulator of NF-$\kappa$B signalling. The hub gene of MDD-downregulated module ME1 is MARCH7, a ubiquitinase that negatively regulates T-cell proliferation (21) (**Figure 3, Table S5**). Notably, the top GO pathway for ME2 (UBXN1-related module) is SCF-dependent proteasomal ubiquitin-dependent protein catabolic process, linking this co-expression module to the pathway-level finding of decreased SCF-related degradation of the cyclin dependent kinase inhibitors in MDD.

The consensus modules identified in females and males were also investigated for their relationship with current smoking status and BMI. Associations between consensus module eigengenes and BMI were tested using Spearman correlation within each dataset, while associations with current smoking status were assessed using Student's t-tests. Two female modules associated with MDD (ME7 and ME4) were significantly associated with BMI in two of the three datasets for which BMI was available (**Figure S11**) (BIODEP, ME7: rho=0.17, p=0.075; ME4: rho=0.085, p=0.377; dbGaP, ME7: rho=0.11, p=2.33e-04; ME4: rho=0.13, p=1.12e-05; Mostafavi, ME7: rho=0.18, p=6.67e-04; ME4: rho=0.17, p=1.65e-03). No male modules were associated with BMI in >1 dataset at unadjusted P<0.05. No male or female modules were associated with smoking status in >1 dataset at unadjusted P<0.05 (**Table S7**).
